# Rotational Percutaneous Mechanical Thrombectomy for Acute and Subacute Limb Ischaemia: a Systematic Review and Proportional Meta-analysis

**DOI:** 10.64898/2026.01.06.26343519

**Authors:** Benedict Stanberry, Koen Deloose, Yann Gouëffic, Gilles Goyault, Giacomo Isernia, Ralph Jackson, Nils Kucher, Michael Lichtenberg, James McCaslin, Bruno Migliara, Mark Portou, Christos Rammos, Caren Randon, Refaat Salman, Marc Sirvent

## Abstract

**Objective:** Rotational percutaneous mechanical thrombectomy with adjunctive angioplasty, stenting or limited thrombolysis (rPMT+) has become commonplace in the treatment of acute and subacute limb ischaemia. This systematic review and proportional meta-analysis synthesises the available evidence on its safety and efficacy.

**Data Sources:** MEDLINE, Embase and the Cochrane Library. ≥

**Review Methods:** We searched for studies (≥10 patients) published since 1 January 2012 (PROSPERO protocol CRD420251015846). Risk of bias was assessed using RoBANS 2. Proportional meta-analysis maximised representative sample sizes by incorporating both single-arm and comparative studies. GRADE evidence profiles were developed for each outcome of interest.

**Results:** Twenty-four studies – seven comparative and 17 single arm – containing 2,954 procedures (2,697 rPMT+, 257 controls) met eligibility criteria. There were no randomised controlled trials. The pooled technical success (TS) rate for rPMT+ was 98% (95% CI: 97-100%, *p*<0.001). Amputation-free survival (AFS) was 96% (95% CI: 93-98%, *p*<0.001). Twelve-month primary patency (PP) was 68% (95% CI: 55-79%, *p*<0.001). The pooled rate of freedom from clinically-driven target lesion revascularisation (fCDTLR) was 85% (95% CI: 80-90%, *p*<0.001). No significant differences existed between acute, subacute and mixed subgroups. Procedure-related mortality was 0.5% (11 deaths in 2,086 procedures from 17 studies) with most deaths occurring after adjunctive catheter-directed thrombolysis. Major adverse events occurred in 4% of procedures (95% CI: 1-8%, *p*<0.001) and distal embolization in 8% (95% CI: 5-11%, *p*<0.001). rPMT+ reduced length of stay by 1.7 days versus alternatives (95% CI: 3.3-0.1 days, *p*<0.05). All outcomes demonstrated heterogeneity. GRADE assessment rated evidence certainty as low to very low.

**Conclusion:** rPMT+ delivers consistently high TS and AFS rates, promising 12-month fCDTLR, acceptable PP, low procedural risk and shorter hospital stays than comparators. The consistency of favourable outcomes supports its use as a safe and effective treatment in appropriately selected ALI and SLI patients.

**WHAT THIS PAPER ADDS:** This systematic review and proportional meta-analysis synthesises 24 single-arm and comparative non randomised studies encompassing nearly 3,000 procedures to provide a comprehensive assessment of the safety and efficacy of rotational percutaneous mechanical thrombectomy in patients with acute and subacute lower limb ischaemia. It demonstrates that, when used for initial revascularisation prior to adjunctive angioplasty, stenting or limited thrombolysis, this endovascular approach delivers consistently high rates of technical success and amputation-free survival with notably high rates of freedom from clinically-driven target lesion revascularisation, acceptable primary patency, low procedural risk and significantly shorter hospital stays.

## INTRODUCTION

Acute limb ischaemia (ALI) is a vascular emergency characterised by a sudden decrease in arterial perfusion, present for ≤14 days, that threatens limb viability and therefore requires urgent evaluation and management.^1^ The most common aetiologies for nontraumatic ALI are acute thrombotic occlusion of a pre-existing stenotic arterial segment or embolus.^2^ Notwithstanding significant advances in its management, it has a poor prognosis: 30-day amputation rates of 10-30% and mortality rates of 15-20%.^3^ Use of the term “subacute limb ischaemia” (SLI) persists, in both clinical practice and peer-reviewed literature, to describe clinical presentations with symptom duration >14 days whose severity and lesion characteristics warrant treatment using the same modalities as are indicated for ALI.^4^

ALI is typically managed through either open surgery (OS) or, more commonly, by percutaneous catheter-directed thrombolysis (CDT).^5,6^ Pooled trial evidence comparing OS to CDT has not reported significant differences in overall death or amputation rates.^7,8^ However, the higher perioperative mortality associated with OS – particularly among elderly and frail patients – has led to CDT becoming the favoured first-line treatment despite its long infusion times and the need for close monitoring in an intensive care unit due to the increased risk of haemorrhage.^9^ Given these risks, percutaneous mechanical thrombectomy (PMT) devices with different mechanisms for thrombus clearance have been gaining popularity.^10^

Rotarex S ™ (BD, Tempe, AZ, USA) is a widely used example of this class of devices. It uses a rotating mechanism to actively modify, excise and aspirate thrombus from peripheral arteries and is indicated for both native blood vessels, native or artificial bypasses and vessels fitted with stents or stent-grafts.^11^ The device is yet to be evaluated in a randomised controlled trial (RCT) and although, over the last two decades, it has been the subject of numerous non-randomised studies of interventions (NRSIs) relatively few of these have been comparative.^12,13^ It was included in a systematic review by Ontario Health that appraised multiple PMT devices and excluded single-arm studies.^14^ Placing such a constraint on the data available for synthesis may have compromised the reliability of the resulting pooled effect summaries for the review’s rotational subgroup, of which Rotarex was the only member. The aim of this review is therefore to inform decision-making by using proportional meta-analyses to generate single summary estimates of the device’s efficacy and safety in the treatment of ALI and SLI by optimally incorporating both single-arm NRSIs and arms from comparative studies to maximise representative sample sizes.^15^

## METHODS

A systematic review and proportional meta-analyses were conducted according to the Preferred Reporting Items for Systematic Reviews and Meta-Analyses (PRISMA) guideline (Supplementary Table S1).^16^ The protocol was prospectively registered in the PROSPERO database (CRD420251015846).^17^

### Eligibility criteria

The full inclusion and exclusion criteria for this review are summarised in PICOS (population, intervention, comparison, outcome, study design) format in Table 1. We included all full text studies of any design with ≥10 patients in any language and from any setting reporting on outcomes of using Rotarex to perform rotational percutaneous mechanical thrombectomy to treat ALI and/or SLI, whether with or without adjunctive angioplasty, stenting or limited thrombolysis (rPMT +). A *post hoc* decision was made to exclude studies published before 1 January 2012. Since the device is indicated for use in peripheral arteries, treatment of deep vein thrombosis was outside the scope of this review. We also excluded children ( < 18 years) and patients with in-stent restenosis or chronic limb-threating ischaemia (symptomatic for > 90 days) without acute or subacute presentation. Separate syntheses were carried out for each outcome of interest with additional sub-group analyses.

**Table 1.**
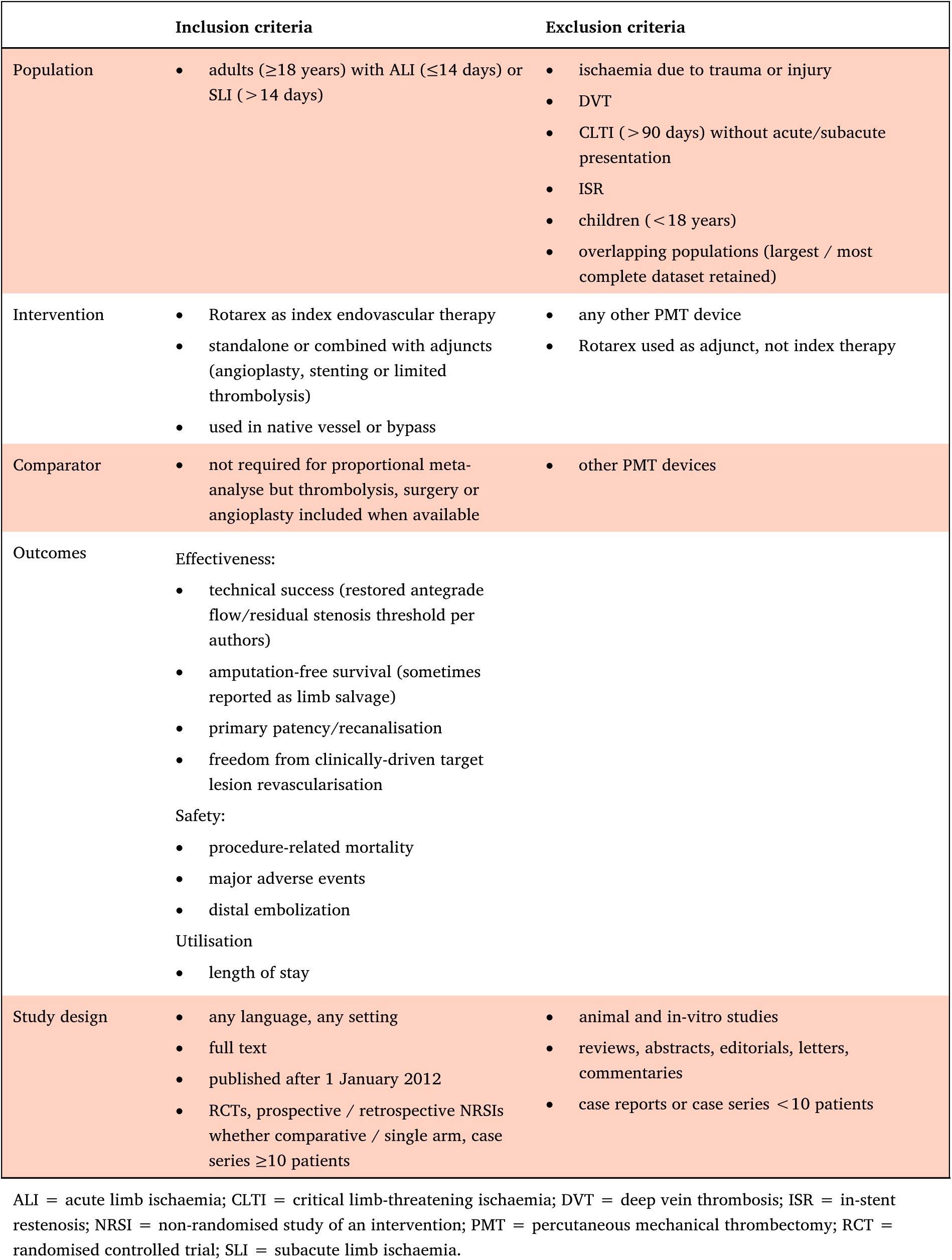
Full inclusion and exclusion criteria for study eligibility.

|  | Inclusion criteria | Exclusion criteria |
| --- | --- | --- |
| Population | adults ($\geq 18$ years) with ALI ($\leq 14$ days) or SLI ($> 14$ days) | ischaemia due to trauma or injury<br>DVT<br>CLTI ($> 90$ days) without acute/subacute presentation<br>ISR<br>children ($< 18$ years)<br>overlapping populations (largest / most complete dataset retained) |
| Intervention | Rotarex as index endovascular therapy<br>standalone or combined with adjuncts (angioplasty, stenting or limited thrombolysis)<br>used in native vessel or bypass | any other PMT device<br>Rotarex used as adjunct, not index therapy |
| Comparator | not required for proportional meta-analyse but thrombolysis, surgery or angioplasty included when available | other PMT devices |
| Outcomes | Effectiveness:<br>technical success (restored antegrade flow/residual stenosis threshold per authors)<br>amputation-free survival (sometimes reported as limb salvage)<br>primary patency/recanalisation<br>freedom from clinically-driven target lesion revascularisation<br>Safety:<br>procedure-related mortality<br>major adverse events<br>distal embolization<br>Utilisation<br>length of stay |  |
| Study design | any language, any setting<br>full text<br>published after 1 January 2012<br>RCTs, prospective / retrospective NRSIs whether comparative / single arm, case series $\geq 10$ patients | animal and in-vitro studies<br>reviews, abstracts, editorials, letters, commentaries<br>case reports or case series $< 10$ patients |

ALI = acute limb ischaemia; CLTI = critical limb-threatening ischaemia; DVT = deep vein thrombosis; ISR = in-stent restenosis; NRSI = non-randomised study of an intervention; PMT = percutaneous mechanical thrombectomy; RCT = randomised controlled trial; SLI = subacute limb ischaemia.

### Information sources, search strategy and selection process

A comprehensive search of MEDLINE, Embase and the Cochrane Library was conducted via the Dialog platform by an information specialist using a pre-specified search strategy (Supplementary Table S2). Reference lists of included studies were checked for additional relevant studies. The main search was carried out on 7 November 2024 and a final search for new studies took place on 1 July 2025.

For MEDLINE and Embase, search strategies were devised using three main concepts: (1) the medical device “Rotarex” or “rotational thrombectomy” or “mechanical thrombectomy” to describe the class of device, (2) alternative terms for CDT, such as “thrombolysis” and “fibrinolysis”, and for the class of drugs used in this treatment, and (3) generic terms used to describe limb ischaemia as the indication that focus on the anatomic aspect of the condition. Where available, both thesaurus terms and textwords were identified to describe each concept. Abbreviations such as ALI were not used due to their potential to retrieve irrelevant references. Two reviewers independently screened the titles and abstracts based on the inclusion and exclusion criteria, followed by full text screening. Disagreements were resolved by consensus. All screening was done manually without the use of artificial intelligence. Authors of included studies were not contacted.

### Data collection process, data items and effect measures

Two independent reviewers used a standardised form to extract data on study characteristics (author, publication year, country), study type (study period, design and size, data source and collection), participant information (cohort sizes, mean age, intervention, comparators, follow-up period), lesion characteristics (occlusion age, type, location and length) and the reported primary and secondary endpoints. The outcomes of interest were pre-specified by a 14-member clinical advisory panel. Four measures of effectiveness were sought: technical success (TS), amputation-free survival (AFS), primary patency (PP) and freedom from clinically-driven target lesion revascularisation (fCDTLR). We sought three measures of safety: procedure-related mortality (PRM), major adverse events (MAE) and distal embolisation (DE) and one measure of utilisation: length of stay (LOS).

### Study risk of bias, reporting bias and outcome certainty assessment

Two reviewers independently assessed the risk of bias of individual studies using the recently revised RoBANS 2 tool.^18^ Though less precise than Cochrane’s ROBINS-I ^19^ tool, RoBANS 2 is less time-consuming to use effectively.^20,21^ The risk of bias of each study was summarised in tabular form to include assessments of its prejudicial potential due to bias in selection, comparability, outcome measurement and reporting. Disagreements were resolved by consensus. Evidence profiles were developed for each outcome using the Grading of Recommendations, Assessment, Development and Evaluations (GRADE) approach.^22^

### Synthesis methods

Given our expectation that most included studies would contain single-arm, non-comparative datasets and our need to determine the pooled prevalence of each outcome across diverse settings, we did not anticipate that standard meta-analyses would be statistically feasible. Meta-analyses of proportions were therefore planned in which variance instability and between-study heterogeneity were addressed using the Freeman-Tukey double arcsine transformation.^15,23^ Effect size and sampling variances were then pooled with random effect modelling using Stata version 18 (StataCorp LLC, College Station, Texas, USA). Between-study variance and heterogeneity were assessed by Q-test and I^2^ statistic with corresponding 95% confidence intervals (CI). Summary group proportion forest plots were created for visual inspection. We planned to conduct exploratory subgroup analysis to investigate the impact of independent variables on outcomes where feasible and appropriate. Sensitivity analysis was planned to assess the robustness of the performed meta-analyses.

## RESULTS

### Study selection

Our search strategy identified 351 publications (Fig. 1). After removal of 90 duplicates, the titles and abstracts of the remaining 261 records were screened against our inclusion and exclusion criteria. Of these, 211 studies were excluded, including 9 studies focused on femoropopliteal in-stent restenosis which the authors agreed should be the subject of a separate review. After screening the full text of 50 studies a further 26 were excluded, leaving 24 studies eligible for inclusion.

**Figure 1.**
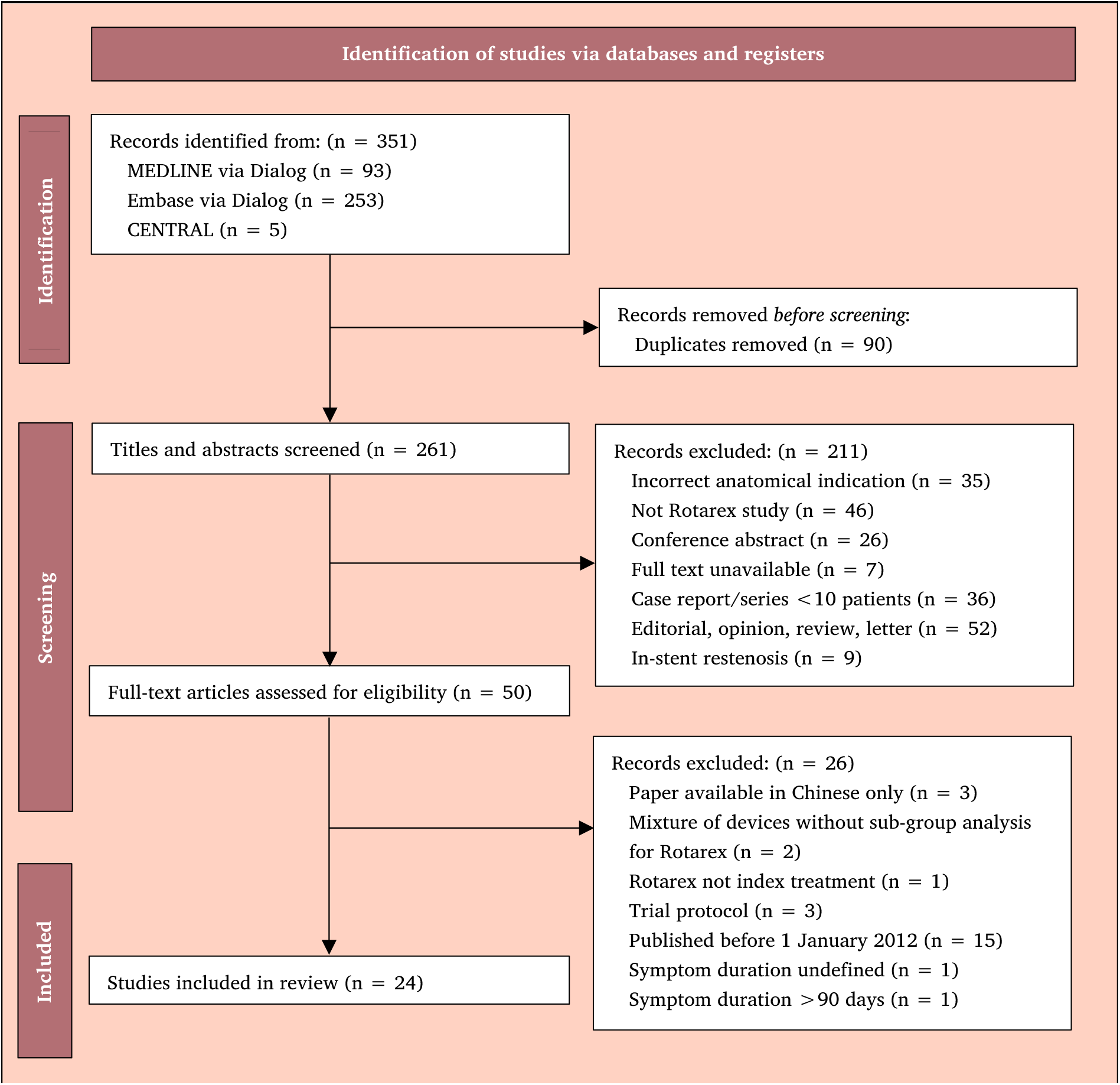
Preferred Reporting Items for Systematic Reviews and Meta-analyses (PRISMA) flow diagram for studies reporting on outcomes of rPMT+ for acute and/or subacute lower limb ischaemia. The last search was performed on 1 July 2025.

### Study and procedure characteristics

The characteristics of the included studies and procedures are summarised in Table 2. Twenty-four studies – of which seven were comparative and 17 were single arm – met the eligibility criteria. There were no RCTs. Three studies were conducted prospectively, while 21 were retrospective. Nine studies were from Germany, nine from China, three from the Czech Republic and one from Italy, Poland, and Taiwan respectively. Study time periods spanned 2005-2022 and reflected contemporary vascular and endovascular treatments options for ALI and SLI. The majority of studies (n = 20) were from single institutions and four were multi-centre. Among the seven comparative studies CDT was the most frequently used control arm (n = 4), followed by surgery (n = 2) and drug-coated balloon (DCB) (n = 1).

**Table 2.** Characteristics of studies and procedures included in the systematic review and proportional meta-analyses reporting on outcomes of rPMT+ for acute and/or subacute lower limb ischaemia.

|  |  |  |  |  |  |  |  |  |  |  |
| --- | --- | --- | --- | --- | --- | --- | --- | --- | --- | --- |
| Author, year, country | Study period | Study design | Data source | Data collection | Ischemia type | rPMT + adjuncts | Control | Procedures – n(%) |  |  |
|  |  |  |  |  |  |  |  | Total | rPMT | Control |
| Li, 2023<br>China <sup>24</sup> | 2018 – 2022 | Single arm | Single centre | Retrospective | Acute | Rotarex + PTA, stent, CDT | — | 17 | 17 (100%) | — |
| Fan, 2023<br>China <sup>25</sup> | 2018 – 2020 | Comparative non-random | Multi-centre | Retrospective | Subacute | Rotarex + DCB, stent | DCB | 74 | 35 (47%) | 39 (53%) |
| Artzner, 2022<br>Germany <sup>26</sup> | 2010 – 2020 | Single arm | Single centre | Retrospective | Acute (n = 267)<br>Subacute (n = 116)<br>Unknown (n = 14) | Rotarex + PTA, DCB, stent, CDT | — | 397 | 397 (100%) | — |
| Zhao, 2022<br>China <sup>27</sup> | 2018 – 2021 | Comparative non-random | Single centre | Retrospective | Acute | Rotarex + PTA, stent | OS | 64 | 32 (50%) | 32 (50%) |
| Yang, 2022<br>China <sup>28</sup> | 2010 – 2020 | Single arm | Single centre | Retrospective | Acute | Rotarex + PTA, stent | — | 38 | 38 (100%) | — |
| Wang, 2022<br>Taiwan <sup>29</sup> | 2016 – 2020 | Comparative non-random | Single centre | Prospective | Acute (n = 56)<br>Subacute (n = 38) | Rotarex + CDT, PTA, stent | CDT | 94 | 66 (70%) | 28 (30%) |
| Liu et al, 2022<br>China <sup>30</sup> | 2015 – 2022 | Single arm | Multi-centre | Retrospective | Acute | Rotarex + PTA, stent, CDT | — | 155 | 155 (100%) | — |
| Stahlberg, 2021<br>Germany <sup>31</sup> | 2014 – 2017 | Single arm | Single centre | Retrospective | Acute (n = 55)<br>Subacute (n = 10) | Rotarex + PTA, DCB, stent | — | 65 | 65 (100%) | — |
| Gong, 2021<br>China <sup>32</sup> | 2015 – 2019 | Comparative non-random | Single centre | Retrospective | Acute | Rotarex + CDT, PTA, stent | CDT | 54 | 13 (24%) | 41 (76%) |
| Fluck, 2021<br>Germany <sup>33</sup> | 2009 – 2016 | Single arm | Single centre | Retrospective | Acute (n = 118)<br>Subacute (n = 14) | Rotarex + PTA, stent, PAT | — | 132 | 132 (100%) | — |
| Wang, 2020<br>China <sup>34</sup> | 2016 – 2018 | Single arm | Multi-centre | Retrospective | Acute (n = 43)<br>Subacute (n = 38) | Rotarex + DCB | — | 81 | 81 (100%) | — |
| Migliara, 2020<br>Italy<sup>35</sup> | 2013 – 2018 | Comparative non-random | Single centre | Retrospective | Acute (n = 10)<br>Subacute (n = 15)<br>Chronic (n = 23) | Rotarex + PTA, stent | OS | 48 | 31 (65%) | 17 (35%) |
| Liu, 2019<br>China<sup>36</sup> | 2016 – 2018 | Single arm | Single centre | Retrospective | Acute<br>Subacute | Rotarex + PTA, stent | — | 42 | 42 (100%) | — |
| Liang, 2019<br>China<sup>37</sup> | 2015 – 2017 | Single arm | Multi-centre | Retrospective | Acute | Rotarex + PTA, stent, CDT | — | 117 | 117 (100%) | — |
| Latacz, 2019<br>Poland<sup>38</sup> | 2014 – 2016 | Single arm | Single-centre | Retrospective | Acute (n = 13)<br>Subacute (n = 38) | Rotarex + PTA, DEB, stent | — | 51 | 51 (100%) | — |
| Bulvas, 2019<br>Czech R<sup>39</sup> | 2009 – 2015 | Single-arm | Single centre | Prospective | Acute (n = 202)<br>Subacute (n = 114) | Rotarex + PTA, stent, PAT, CDT | — | 316 | 316 (100%) | — |
| Kronlage, 2017<br>Germany<sup>40</sup> | 2006 – 2015 | Comparative non-random | Single centre | Retrospective | Acute (n = 78)<br>Subacute (n = 124) | Rotarex only (n = 146)<br>Rotarex + CDT (n = 28) | CDT | 202 | 146 (72%)<br>28 (14%) | 28 (14%) |
| Heller, 2017<br>Czech R<sup>41</sup> | 2008 – 2015 | Single arm | Single centre | Retrospective | Acute | Rotarex + PTA, stent, PAT, CDT | — | 120 | 120 (100%) | — |
| Freitas, 2017<br>Germany<sup>42</sup> | 2005 – 2013 | Single arm | Single centre | Retrospective | Acute (n = 211)<br>Subacute (n = 314) | Rotarex + PTA, stent, CDT | — | 525 | 525 (100%) | — |
| Stanek, 2016<br>Czech R<sup>43</sup> | 2007 – 2014 | Single arm | Single centre | Prospective | Acute<br>Subacute | Rotarex + PTA, stent | — | 128 | 128 (100%) | — |
| Scheer, 2015<br>Germany<sup>44</sup> | NR | Single arm | Single centre | Retrospective | Acute (n = 8)<br>Subacute (n = 21) | Rotarex + PTA, stent | — | 29 | 29 (100%) | — |
| Wissgott, 2013<br>Germany<sup>45</sup> | 2008 – 2012 | Single arm | Single centre | Retrospective | Acute (n = 31)<br>Subacute (n = 11) | Rotarex + PTA, stent | — | 42 | 42 (100%) | — |
| Lichtenberg, 2013<br>Germany<sup>46</sup> | 2008 – 2010 | Single arm | Single centre | Retrospective | Acute<br>Subacute | Rotarex + PTA, stent, CDT | — | 22 | 22 (100%) | — |
| Hundt et al,<br>2013<br>Germany <sup>47</sup> | 2007 –<br>2012 | Comparative | Single<br>centre | Retrospective | Acute<br>(n = 75)<br>Subacute<br>(n = 66) | Rotarex +<br>CDT, PTA,<br>stent | CDT | 141 | 69<br>(49%) | 72<br>(51%) |
| **Total** |  |  |  |  |  |  |  | **2,954** | **2,697** | **257** |

CDT = catheter-directed thrombolysis; DCB = drug-coated balloon; DEB = drug-eluting balloon; OS = open surgery; NR = not reported; PAT = percutaneous aspiration thrombectomy; PTA = percutaneous balloon angioplasty; rPMT+ = rotational percutaneous mechanical thrombectomy plus adjuncts.

Overall, 2,954 procedures for ALI and SLI were included, with 2,697 in rPMT + arms and 257 in control arms. Seven studies investigated patients with ALI only,^24,27–28,30,32,37,41^ one study focused exclusively on SLI,^25^ and the remaining 16 studies contained a mixture of both ALI and SLI patients.^26,29,31,33–36,38–40,42–47^ There were a total of 1,732 (58.6%) clearly identified ALI patients and 993 SLI patients (33.6%). Three studies, containing 192 patients (6.5%) did not sort ALI and SLI patients into subgroups.^36,43,46^ In most studies (n = 20) rPMT + was used as index treatment with adjunctive balloon angioplasty and/or stenting performed after bypass revascularisation as needed. Ten studies explicitly stated CDT was used adjunctively in cases of inadequate removal of thrombus load or when a thromboembolism dropped in the infrapopliteal arteries.

### Patient and outcome characteristics

The characteristics of the patients and outcomes in each eligible study are summarised in Table 3. The mean age of the included patients was 69.3 years (range: 61.5-80.0 years). Mean follow-up time was 13.1 months (range: 1-42 months). Fourteen (54.2%) studies reported mean lesion length – which was 20.1 cm (range: 11.0-32.1 cm) for all included patients.

**Table 3.**
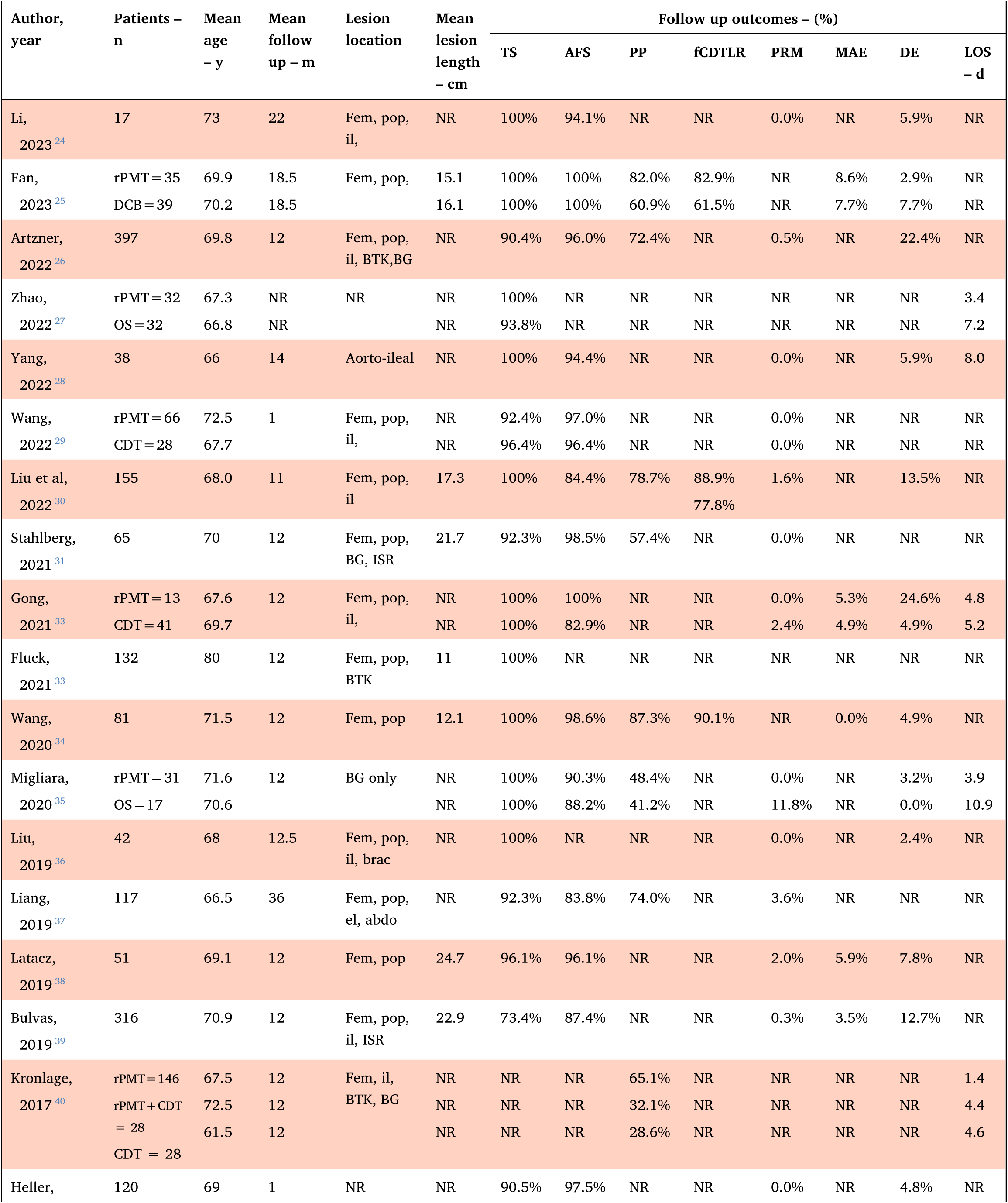
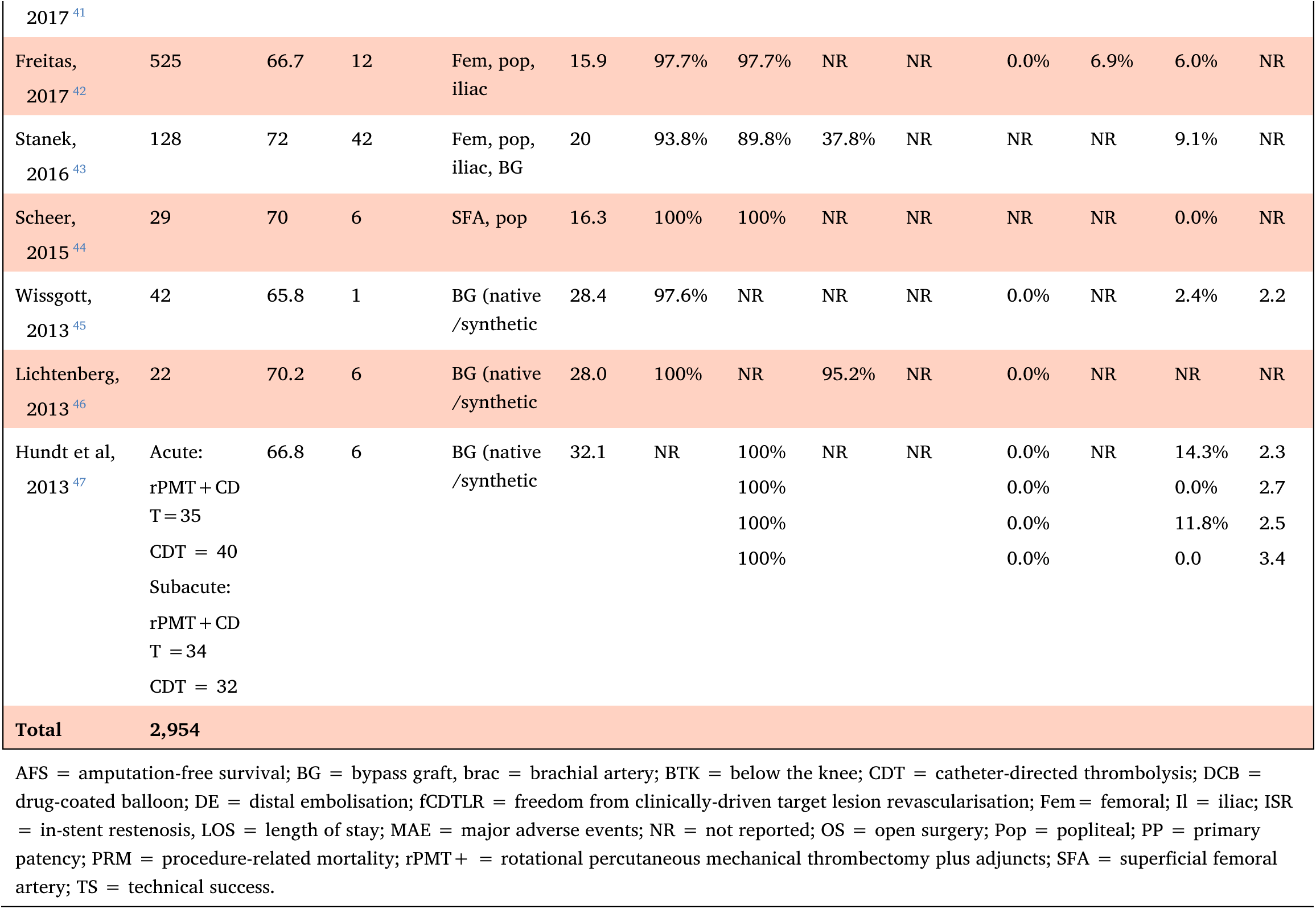
Characteristics of patients and outcomes included in the systematic review and proportional meta-analyses reporting on outcomes of rPMT+ for acute and/or subacute lower limb ischaemia.

The most frequently reported outcome was TS, which was provided in 22 (92%) of the included studies. Eighteen (75%) studies reported AFS. PP was reported in 10 (42%) studies and fCDTLR in 3 (13%) studies. Only 4 (17%) studies reported on all four of the primary endpoints we sought. Only 4 (17%) studies contained all three pre-defined safety outcomes and 3 (13%) studies did not contain any. LOS was reported in 7 (29%) studies.

### Risk of bias in studies

The results of the assessment of bias for each included study are summarised in Supplementary Table S3. The RoBANS 2 criteria evaluated 3 studies to be at moderate risk of bias and 21 studies to have a high risk of bias.

### Technical success

Twenty-two (92%) studies reported the immediate TS rate of which 5 (21%) were comparative.^25,27,29,32,35^ Zhao et al found rPMT + achieved greater success than surgical thrombectomy or resection (100% v 93.8%, *p*=0.162) whereas Migliara et al observed that, in patients with bypass occlusions, both rPMT + and surgery delivered 100% success rates.^27,35^ Wang et al observed that rPMT +CDT achieved slightly poorer TS than CDT alone (92.4% v 96.4%, *p*=0.47) while Gong et al noted both interventions had 100% success in restoring continual arterial patency (*p*=0.584).^29,32^ Comparing rPMT+DCB with DCB alone in a subacute patients, Fan et al saw 100% success rates achieved by both procedures (Table 3).^25^ Twenty-two studies with 2,485 procedures were included in the proportional meta-analysis for TS. The overall pooled success rate for rPMT + was 98% (95% CI: 97-100%, *p*<0.001) with substantial heterogeneity (*I*^2^=80%) (Fig. 2). There were no significant differences between the pooled success rates for ALI patients (99%, 95% CI: 95-100%, *p*<0.001), SLI patients (100%, 95% CI: 95-100%, p<0.001) or mixed ALI/SLI populations (98%, 95% CI: 96-99%, *p*<0.001). The GRADE certainty for the evidence is low, with downgrading for risk of bias and imprecision (Table 4).

**Figure 2.**
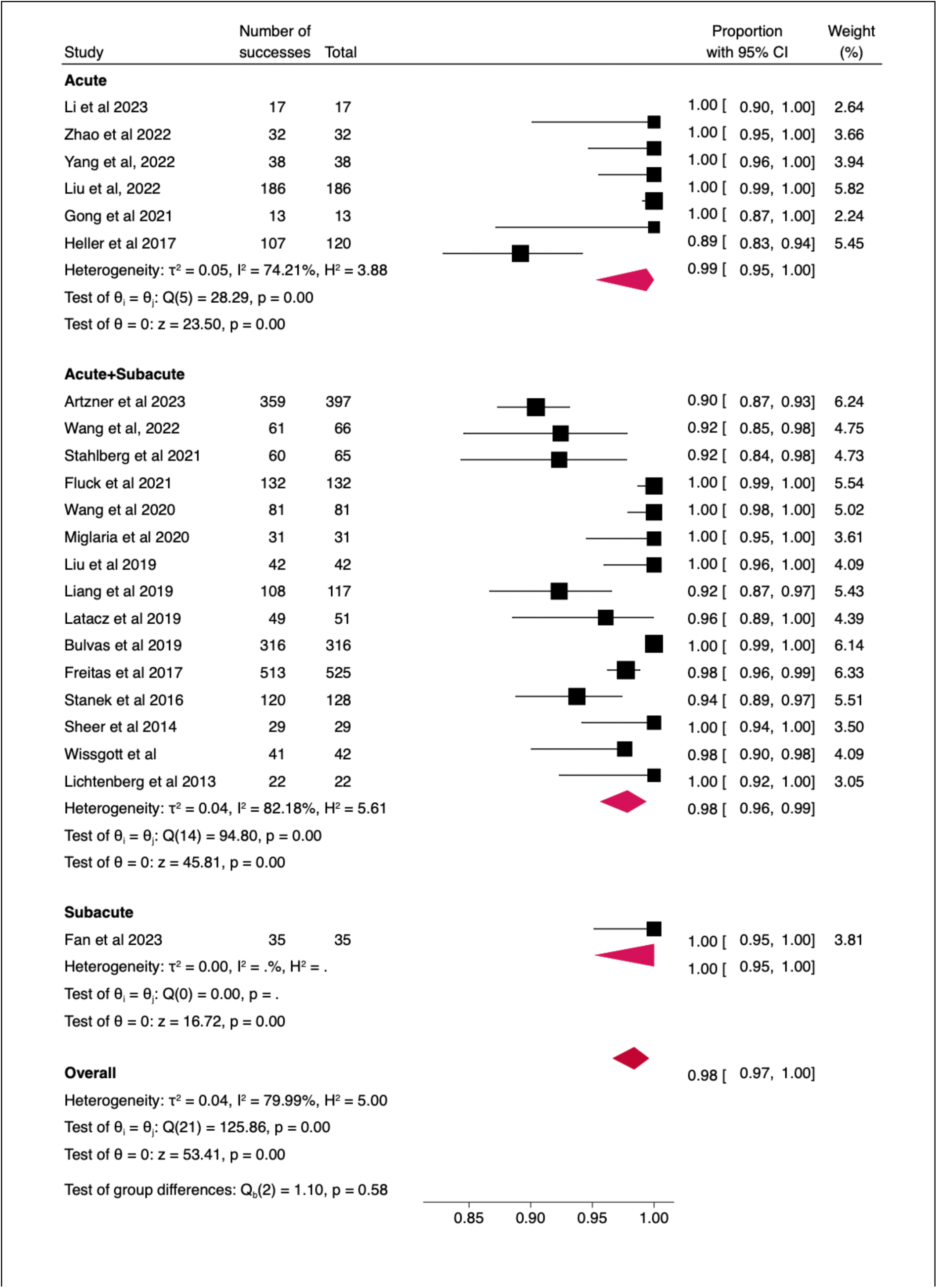
Forest plot of proportional meta-analysis of 22 studies including 2,485 procedures illustrating the pooled technical success rate for rPMT + in acute, acute/subacute and subacute lower limb ischaemia.

**Table 4.**
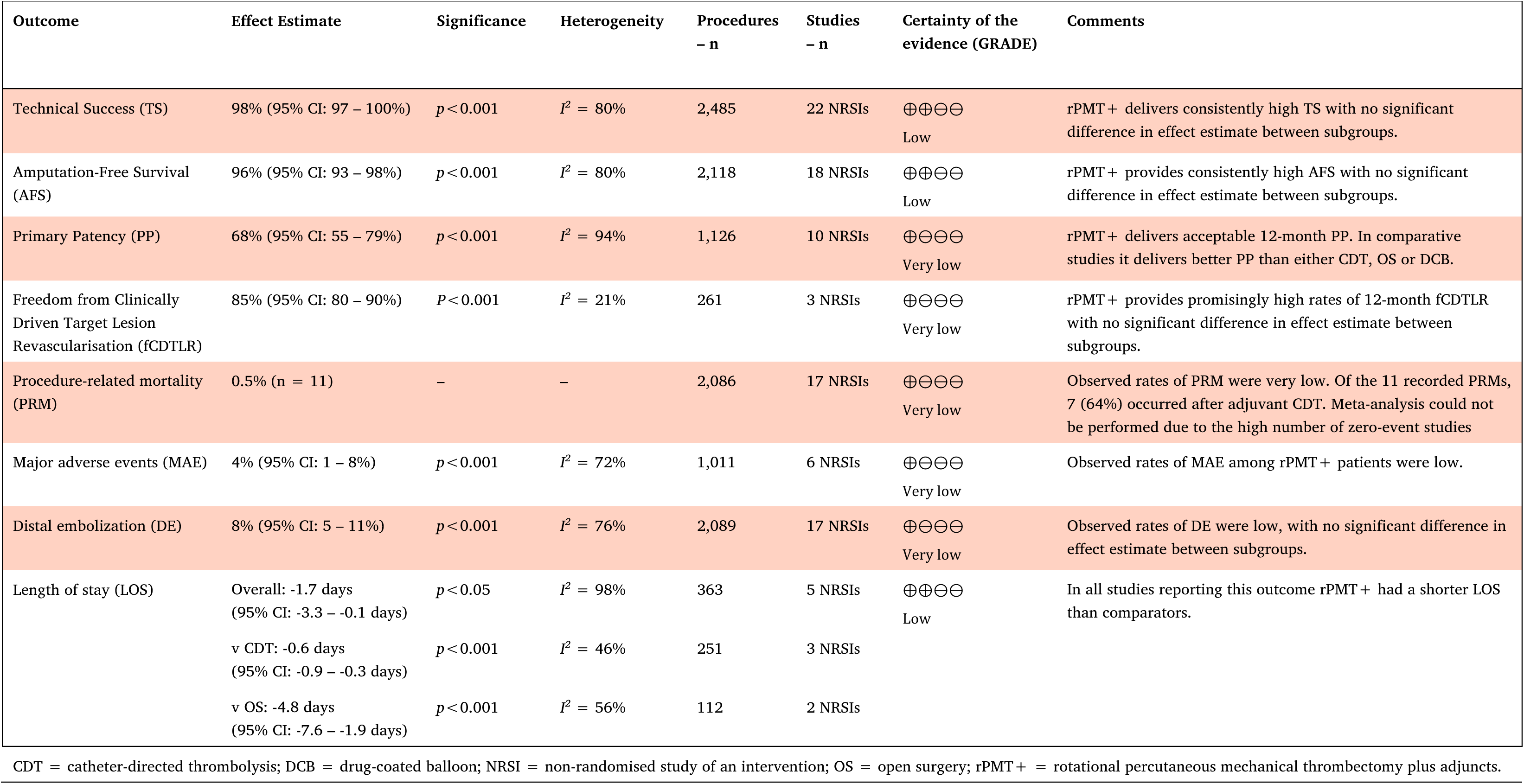
Summary of findings and Grading of Recommendations Assessment, Development and Evaluation (GRADE) assessment for outcomes of rPMT+ for acute and/or subacute lower limb ischaemia.

### Amputation-free survival

Eighteen (75%) studies reported AFS of which five were comparative.^25,29,32,35,47^ Fan et al noted that rPMT +DCB and DCB alone were both associated with 100% AFS at 12 months.^25^ In patients with bypass grafts, Migliara et al saw rPMT + deliver marginally better AFS than surgery (90.3% v 88.2%, *p*=0.832).^35^ Comparing rPMT+CDT to CDT alone, Wang et al observed similar 12-month AFS (97.0% v 96.4%, *p*=0.89) in a mixed acute/subacute population while Hundt et al analysed results for acute and subacute groups separately, recording 100% AFS for both interventions in both groups.^29,47^ On the other hand, Gong et al saw patients receiving rPMT+CDT obtaining slightly better AFS than those receiving CDT alone (89.5% v 82.9%, *p*=0.34).^32^

Eighteen studies that followed up on 2,118 procedures were included in the proportional meta-analysis for AFS. The overall pooled success rate for rPMT+ was 96% (95% CI: 93-98%, *p*<0.001) with substantial heterogeneity (*I^2^*=80%) (Fig. 3). There were no significant differences between the pooled success rates for ALI patients (96%, 95% CI: 90-100%, *p*<0.001), SLI patients (100%, 95% CI: 97-100%, *p* < 0.001) or the mixed ALI/SLI subgroup (95%, 95% CI: 92-98%, *p*<0.001). The GRADE certainty for the evidence is low, with downgrading for risk of bias and imprecision (Table 4).

**Figure 3.**
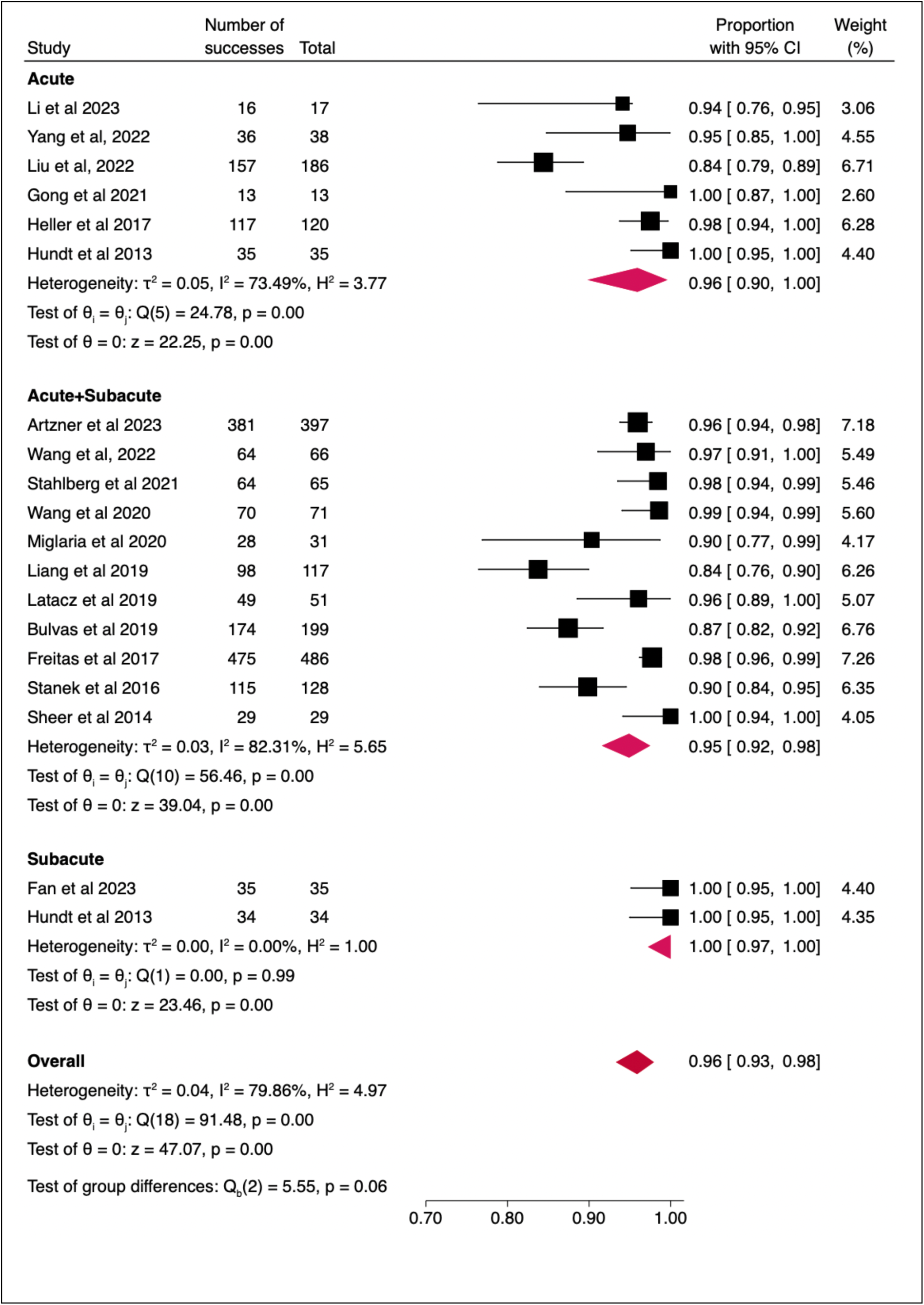
Forest plot of proportional meta-analysis of 18 studies including 2,118 procedures illustrating the pooled amputation-free survival for rPMT + in acute, acute/subacute and subacute lower limb ischaemia.

### Primary patency

PP at 12 months follow-up was calculated in 10 (42%) of the included studies. All studies used the Kaplan-Meier method, except that of Stanek et al that used life tables.^43^ All three of the comparative studies reported rPMT + cohorts achieving better outcomes than comparators.^25,35,40^ Fan et al found rPMT+DCB achieved better patency than DCB alone (82.0% v 60.9%, *p*=0.04).^25^ Migliara et al saw a small patency advantage for rPMT + patients compared to those undergoing surgery (48.4% v 41.2%) and Kronlage observed rPMT alone achieved better PP than either rPMT +CDT or CDT alone (65.1% v 32.1% v 28.6%, *p*<0.0001).^35,40^

Ten studies that followed up on 1,126 procedures were included in the proportional meta-analysis for PP. The overall pooled rate for 12-month PP for rPMT + was 68% (95% CI: 55-79%, *p* < 0.001) with substantial heterogeneity (*I^2^* = 94%) (Fig. 4). The GRADE certainty for the evidence is very low, with downgrading for risk of bias and imprecision (Table 4).

**Figure 4.**
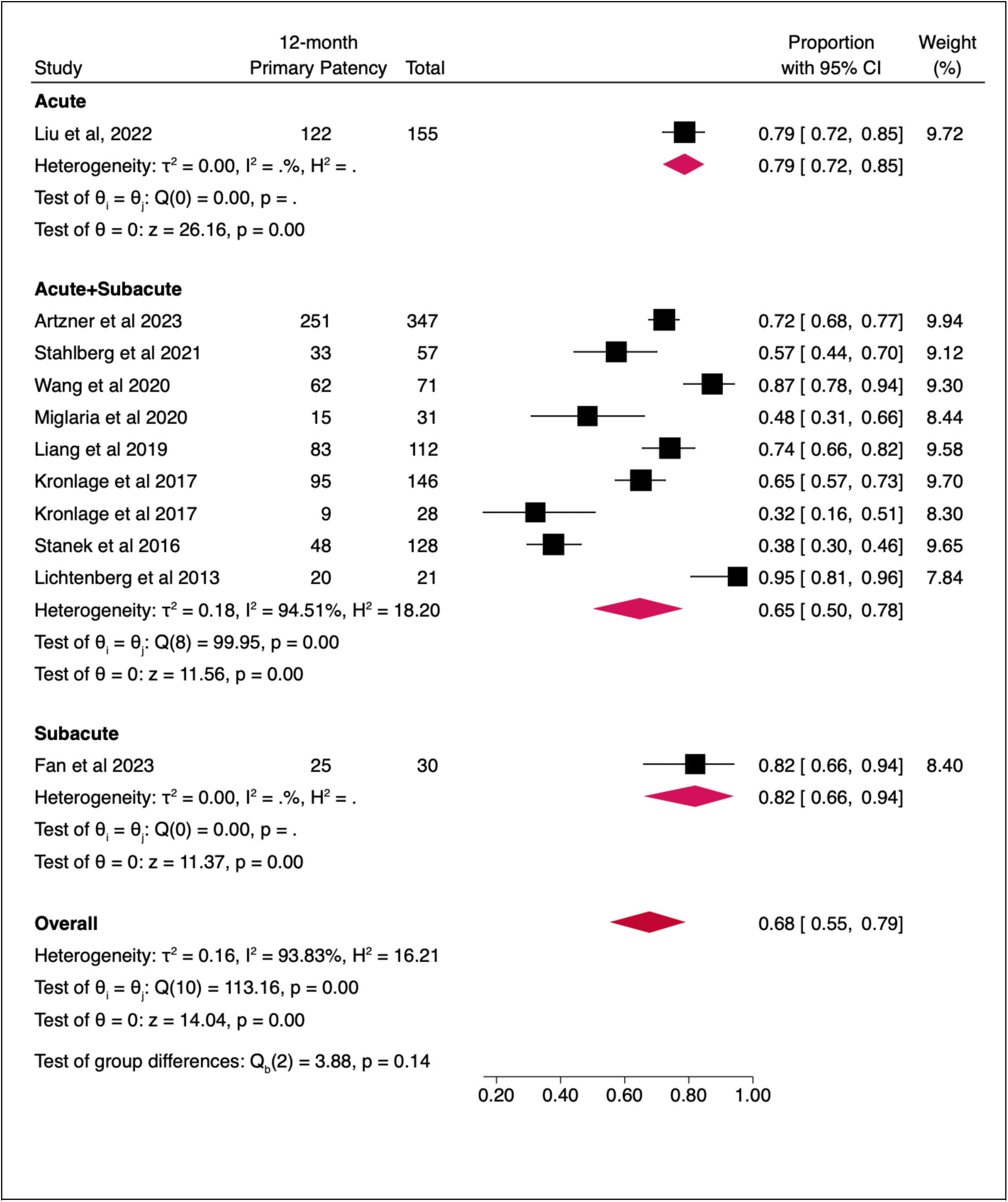
Forest plot of proportional meta-analysis of 10 studies including 1,126 procedures illustrating the pooled 12-month primary patency for rPMT + in acute, acute/subacute and subacute lower limb ischaemia.

### Freedom from clinically-driven target lesion revascularisation

Only three (13%) of the included studies, that included 261 procedures, constructed survival curves for fCDTLR at 12-months follow-up.^25,30,34^ In their comparison of subacute patients, Fan et al observed fCDTLR of 82.9% in the rPMT+DCB group and 61.5% for DCB alone (*p*=0.04).^25^ Liu et al noted fCDTLR differences between different acute aetiologies treated with rPMT+: 88.9% in the embolization group, 77.8% in the thrombosis group and 67.5% in the restenosis group (*p*<0.01).^30^ In a mixed population of acute and subacute patients, Wang et al observed that, one year on from the index procedure, 64 (90%) of 71 patients had not required reintervention.^34^ The overall pooled fCDTLR rate for rPMT + at 12 months was 85% (95% CI: 80-90%, p< 0.001) with low heterogeneity (*I^2^*=21%) (Fig. 5). The GRADE certainty for the evidence is very low, with downgrading for risk of bias and imprecision (Table 4).

**Figure 5.**
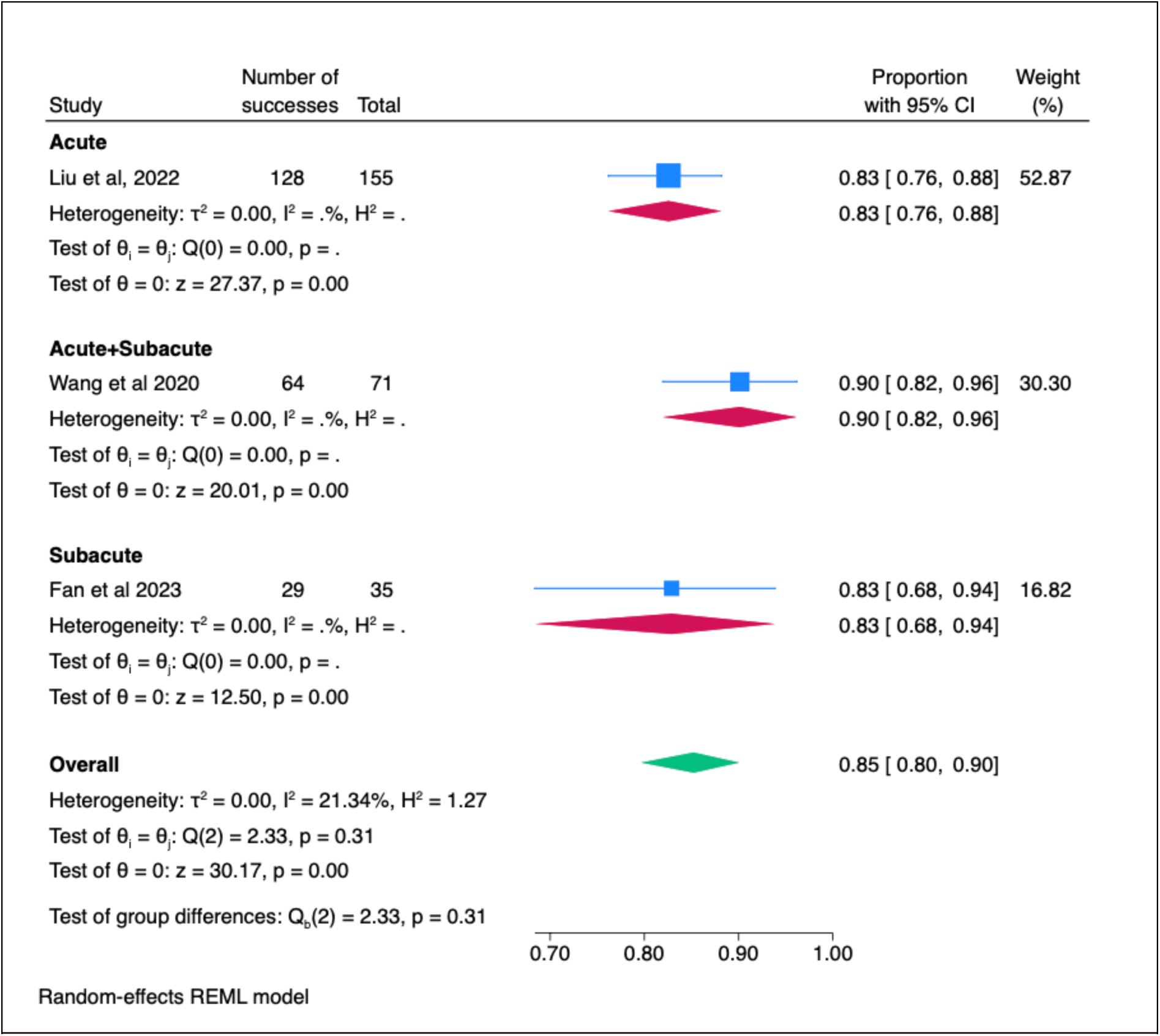
Forest plot of proportional meta-analysis of 3 studies including 261 procedures illustrating the pooled 12-month freedom from clinically-driven target lesion revascularisation rate for rPMT + in acute, acute/subacute and subacute lower limb ischaemia.

### Procedure-related mortality

Seventeen studies (71%) that followed up on 2,244 procedures reported on 30-day PRM or provided sufficient detail to distinguish PRM from non-procedure-related deaths. In 12 (71%) of these studies there were no deaths among patients treated with rPMT +.^24,28–29,31–32,35–36,41–42,45–47^ Two of the studies with zero mortalities among rPMT + patients reported control group mortalities. Miglaria et al reported two (11.8%) procedure-related deaths in the surgical arm of their study while Gong et al reported one (2.4%) PRM from intracranial haemorrhage in their study’s CDT-only arm.^32,35^ PRMs occurred among rPMT + patients in five studies.^26,30,37–39^ In four of them, patients died from haemorrhagic stroke after adjuvant CDT.^26,30,38–39^ In the fifth study, that of Laing et al, four (3.6%) deaths were observed after the rPMT + procedure: one pulmonary infection in a patient with acute arterial thrombosis, one myocardial infarction in a patient with acute arterial embolism and two heart failures, one of which was a patient with stent/graft thrombosis.^37^ A total of 11 (0.5%) PRMs occurred over 2,086 reported rPMT+ procedures. However, there were too many zero-event studies to perform proportional meta-analysis for this outcome.

### Major adverse events

Six (25%) studies covering 1,011 procedures reported the incidence of MAEs.^25,33–34,38–39,41^ Five of these studies used the Society of Interventional Radiology’s definition of major complications.^48^ Freitas et al observed 36 MAEs (6.9%) occurring during the hospital stay of the 525 patients in their study.^42^ At 30-days, the rPMT + arm of Gong et al had a marginally worse 30-day MAE rate than the CDT-only arm (6.3% v 4.9%, *p* = 1.000).^33^ At 12-months follow-up, Wang et al did not record any major complications whereas Latacz et al recorded 3 (5.9%) MAEs among 51 patients.^34,38^ Fan et al reported a slightly higher 18-month MAE rate in their rPMT +DCB group than in their DCB-only group (8.6% v 7.7%, *p* = 0.61).^25^ One study, that of Bulvas et al, used the Society for Vascular Surgery’s criteria for major adverse limb events (MALE).^49^ Among their early postoperative outcomes they noted 11 (3.5%) serious sequelae, 5 (2.2%) of which were major bleeding following adjunctive infrapopliteal thrombolysis. At 12 months they noted patients treated for emboli enjoyed significantly fewer MALEs than those treated for thromboses (3% vs 25%).^39^ The overall pooled MAE rate for rPMT + was 4% (95% CI: 1-8%, p *<* 0.001) with substantial heterogeneity (*I^2^* = 72%) (Fig. 6). The GRADE certainty for the evidence is very low, with downgrading for risk of bias and imprecision (Table 4).

**Figure 6.**
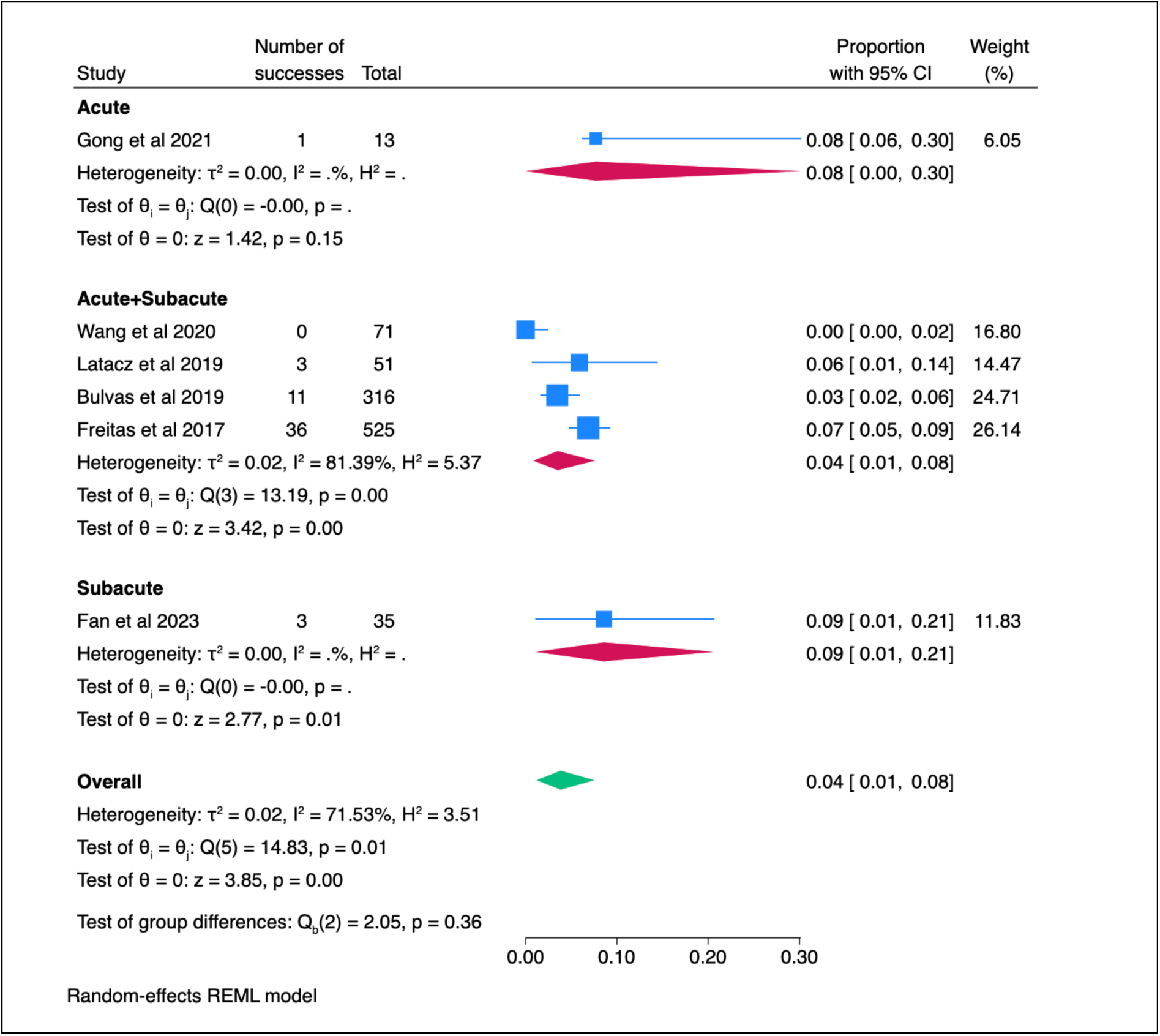
Forest plot of proportional meta-analysis of 6 studies including 1,011 procedures illustrating the pooled major adverse event rate for rPMT + in acute, acute/subacute and subacute lower limb ischaemia.

### Distal embolisation

DE was reported to be a minor procedure-related complication in 17 (71%) of the included studies, four of which were comparative. Migliara et al reported that DE was experienced by 1 (3.2%) of the 31 patients treated for bypass graft occlusions with rPMT + but not by any patient undergoing OS.^35^ Hundt et al observed 5 (14.3%) acute patients and 4 (11.8%) subacute patients treated with rPMT + requiring aspiration and local thrombolysis to manage a DE (which patients in the CDT-only arm of this study did not experience).^47^ Gong et al managed DE in 5 (38.5%) of their rPMT + patients and 2 (4.9%) of their CDT-treated patients.^33^ In their comparison of rPMT +DCB with DCB alone, Fan et al recorded a lower rate of DE in rPMT +DCB patients that those treated with DCB alone (2.9% v 7.7%).^25^

Seventeen studies with 2,089 procedures were included in the proportional meta-analysis for DE. The overall pooled rate of DE for rPMT + was 8% (95% CI: 5% – 11%, *p*<0.001) with substantial heterogeneity (*I^2^* = 76%) (Fig. 7). There were no significant differences between the pooled success rates for ALI patients (9%, 95% CI: 5-15%, *p*<0.001), SLI patients (7%, 95% CI: 0-18%, *p* = 0.01) or mixed ALI/SLI populations (7%, 95% CI: 4-11%, *p*<0.001). Sensitivity analysis by excluding one study at a time did not show any single study heavily influencing the pooled DE rate. The GRADE certainty for the evidence is very low, with downgrading for risk of bias and imprecision (Table 4).

**Figure 7.**
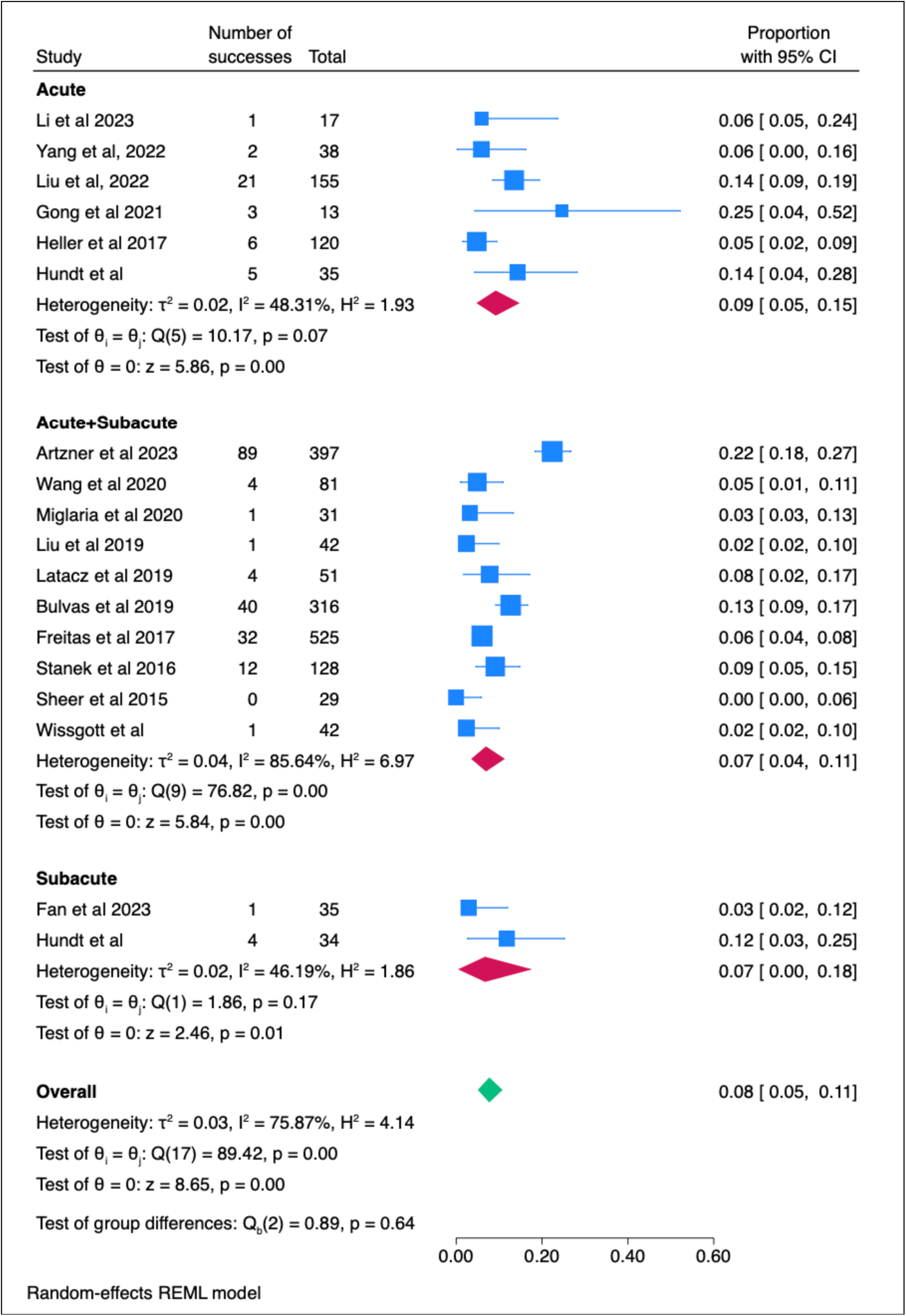
Forest plot of proportional meta-analysis of 17 studies including 2,089 procedures illustrating the pooled distal embolisation rate for rPMT + in acute, acute/subacute and subacute lower limb ischaemia.

### Length of stay

In all 5 (21%) of the comparative studies that measured this endpoint, containing a total of 363 patients, those in rPMT + groups had a shorter LOS in hospital compared to those in control groups.^27,32,35,40,47^ The overall pooled mean difference in LOS between rPMT + and comparators was 1.65 days (95% CI: 0.05-3.25, p< 0.05). The difference in LOS between rPMT + and CDT was 0.60 days (95% CI: 0.26-0.94, *p*<0.001) and between rPMT + and surgery was 4.76 days (95% CI: 1.91-7.61, *p*<0.001) (Fig. 8). Overall heterogeniety was high (*I^2^*=98%) but was moderate in the CDT (*I^2^*=46%) and surgery (*I^2^*=56%) subgroups. The GRADE certainty for the evidence is low, with downgrading for risk of bias (Table 4).

**Figure 8.**
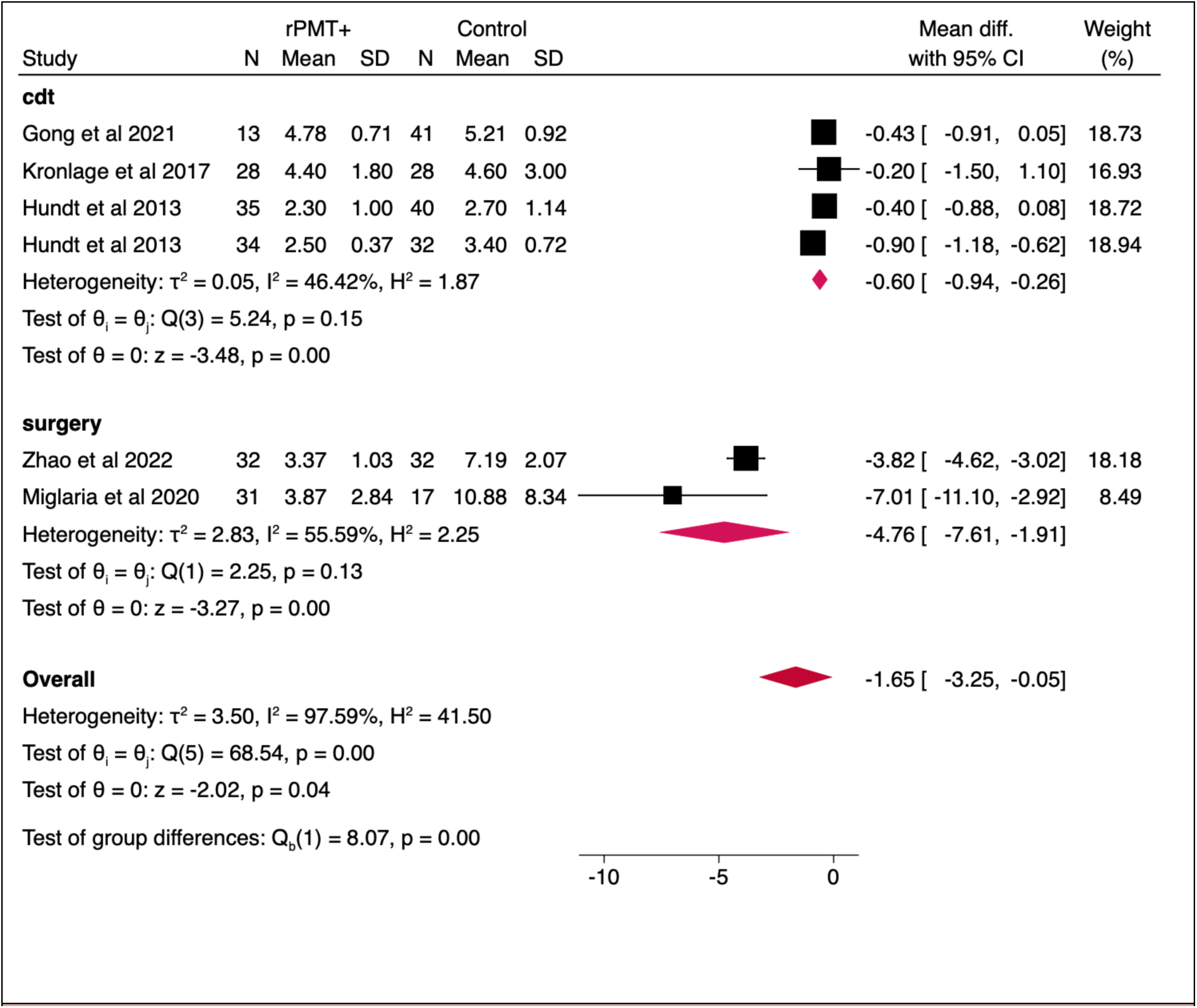
Forest plot of meta-analysis of 5 studies comparing the mean difference in lengths of stay between 363 patients treated for acute, acute/subacute and subacute lower limb ischaemia with rPMT, CDT or surgery.

## DISCUSSION

This systematic review and proportional meta-analysis provides a thorough and comprehensive synthesis of the safety and efficacy outcomes achieved across 24 real-world observational studies conducted over a 17-year total study period and encompassing almost three thousand rPMT + procedures in which the Rotarex rotational percutaneous mechanical thrombectomy system was used for initial vessel preparation and/or revascularisation prior to adjunctive angioplasty and/or stenting and/or limited thrombolysis in patients with ALI and/or SLI.

The achievement of consistently high TS in multiple independent clinical settings may reflect the advantages of active mechanical fragmentation over the passive fragmentation of rheolytic devices or the enzymatic clot-dissolving action of CDT.^50^ The 98% pooled TS rate resulting from our meta-analysis compares very favourably with the 75% pooled mean angiographic success rate for CDT obtained by the systematic review of Ebben et al and the 74% rate for OS reported by the STILE trial.^51,52^ The pooled 96% 12-month AFS rate suggests that inceptive debulking supports durable limb salvage outcomes that exceed the 65% one-year AFS rates reported for open surgical revascularisation by the TOPAS trial and the 75% rates reported for CDT by both the TOPAS and Rochester trials.^53,54^ Lack of standardised reporting of PP for OS makes direct comparisons of this outcome challenging. Although lower at 68%, the 12-month PP achieved by rPMT + is similar to the 70% rate for OS reported by de Donato et al at 24-months and much superior to the 48% PP for CDT alone obtained by the meta-analysis of Doelare et al.^55,56^ The fact that the three studies that measured 12-month fCDTLR reported that this outcome exceeded 80% at one year suggests that the majority of patients treated with rPMT + maintain patency without requiring reintervention.

At 0.5%, PRM was very low and most commonly caused by haemorrhagic stroke occurring after adjuvant thrombolysis, demonstrating that rPMT + provides a compelling safety advantage when compared to the composite 30-day mortality rates associated with OS (12%) and CDT (7%).^57^ The low MAE rate of 4% also compares very favourably with OS and CDT, both of which carry well-documented bleeding risks.^51^ DE was relatively uncommon. The 8% pooled estimate for this complication is subject to considerable definitional heterogeneity such that even minor events that were managed with successful aspiration or limited adjunctive thrombolysis may have been included. Moreover, the causes of the DEs in our pooled estimate are likely to be masked by extensive use of adjunctive procedures such as balloon angioplasty during which residual thrombi may have been dislodged during balloon post-dilation rather than during the thrombectomy procedure itself.^41^ The reduction in LOS – by approximately five days compared with OS and one day compared with CDT – can provide significant cost-savings and enhance the productivity of vascular and endovascular services.^58^

### Strengths and limitations

In the absence of RCTs this study of rPMT + has used proportional meta-analysis of NRSIs to maximise sample size and analytical power, enabling a rigorous statistical synthesis of the currently available evidence and deriving stable estimates of rates of rare events using a transparent, reproducible method that complies with PRISMA and Cochrane standards.^16,59^ The consistency of favourable outcomes across diverse settings, particularly the high TS and AFS rates, provide reasonable confidence that rPMT+ represents an effective strategy for appropriately selected ALI and SLI patients and justifies its continued inclusion in the European Society of Vascular Surgery’s clinical practice guidelines on the management of ALI.^1^ The low PRM and MAE rates support consideration of rPMT + as a first-line endovascular option in centres with appropriate expertise. However, several important limitations warrant acknowledgement. Given the dominance of retrospective studies (21 of 24) it is highly likely that rPMT + patient selection was strongly biased by factors such as thrombus burden and fitness for surgery. Outcome reporting was inconsistent and incomplete – particularly as regards important primary endpoints such as PP and fCDTLR – and was confounded by considerable definitional heterogeneity. TS thresholds, for instance, ranged from restoration of any antegrade flow to achievement of < 30% residual stenosis. Given the retrospective nature of the evidence, some outcome reporting may have been selective. Despite favourable pooled estimates, substantial heterogeneity was observed across practically all primary and secondary endpoints – reflecting differing symptom severity, lesion characteristics, anatomic locations and vessel types as well as considerable variation in the intensity of adjunctive therapy. Device specifications (6F versus 8F catheters), operator experience and lesion-level characteristics were rarely reported and incompletely described, precluding meaningful subgroup analyses of variables that are likely to affect procedural success and complication rates. Publication bias cannot be excluded. These limitations resulted in low or very low GRADE certainty ratings for all outcomes, reflecting serious concerns about risk of bias and imprecision. Future registry-based studies with mandatory reporting could provide real-world safety and efficacy data at scale.

### Conclusions

rPMT + delivers consistently high pooled TS and AFS rates, promising 12-month fCDTLR, acceptable PP, low procedural risk and shorter hospital stays than either OS or CDT. There was no significant difference in pooled effect estimates between ALI, SLI and mixed ALI/SLI subgroups. The consistency of favourable outcomes supports rPMT + as a safe and effective first-line or hybrid treatment strategy for appropriately selected ALI and SLI patients.

## Data Availability

All data produced in the present study are available upon reasonable request to the authors.

## Acknowledgements

The authors wish to thank Beata Coffey, Information Specialist at the Royal Society of Medicine, for her assistance in developing a search strategy and performing the literature search. We also thank Laura Parker who, in her capacity as second reviewer, independently screened reports, assessed them for eligibility and helped select reports for inclusion by consensus with Benedict Stanberry.

## CRediT Authorship Contribution Statement

**Benedict Stanberry:** Conceptualization, Methodology, Software, Formal analysis, Investigation, Resources, Data Curation, Writing – Original Draft, Writing – Review & Editing, Visualisation, Project administration. **Koen Deloose:** Conceptualisation, Validation, Writing – Review & Editing, Supervision. **Yann Gouëffic:** Conceptualisation, Validation, Writing – Review & Editing, Supervision. **Gilles Goyault:** Conceptualisation, Validation, Writing – Review & Editing, Supervision. **Giacomo Isernia:** Conceptualisation, Validation, Writing – Review & Editing, Supervision. **Ralph Jackson:** Conceptualisation, Validation, Writing – Review & Editing, Supervision. **Nils Kucher:** Conceptualisation, Validation, Writing – Review & Editing, Supervision. **Michael Lichtenberg:** Conceptualisation, Validation, Writing – Review & Editing, Supervision. **James McCaslin:** Conceptualisation, Validation, Writing – Review & Editing, Supervision. **Bruno Migliara:** Conceptualisation, Validation, Writing – Review & Editing, Supervision. **Mark Portou:** Conceptualisation, Validation, Writing – Review & Editing, Supervision. **Christos Rammos:** Conceptualisation, Validation, Writing – Review & Editing, Supervision. **Caren Randon:** Conceptualisation, Validation, Writing – Review & Editing, Supervision. **Refaat Salman:** Conceptualisation, Validation, Writing – Review & Editing, Supervision. **Marc Sirvent:** Conceptualisation, Validation, Writing – Review & Editing, Supervision.

Generative AI and AI-assisted technologies were not used in the preparation of this work.

## References

1 Björck M, Earnshaw J, Acosta S, Bastos Gonçalves F, Cochennec F, Debus E, et al. Editor’s Choice – European Society for Vascular Surgery (ESVS) 2020 Clinical Practice Guidelines on the Management of Acute Limb Ischaemia. European Journal of Vascular and Endovascular Surgery 2020; 59(2): 173–218. 10.1016/j.ejvs.2019.09.006

2 Callum K, Bradbury A. ABC of arterial and venous disease: Acute limb ischaemia. BMJ 2000; 320(7237): 764–767. 10.1136/bmj.320.7237.764.

3 Mannan F, Ugwumba L, El-Sayed T, Saratzis A, Nandhra S, ESTABLISh Research Collaborative and the Vascular and Endovascular Research Network. A survey of contemporary acute lower limb ischaemia management. Journal of Vascular Societies Great Britain and Ireland 2025; 4(3): 144–150. 10.54522/jvsgbi.2025.177

4 Dosluoglu H, Harris L. Endovascular Management of Subacute Lower Extremity Ischemia. Seminars in Vascular Surgery 2008; 21(4): 167–179. 10.1053/j.semvascsurg.2008.11.002

5 Baril D, Patel V, Judelson D, Goodney P, McPhee J, Hevelone N, et al. Outcomes of lower extremity bypass performed for acute limb ischaemia. Journal of Vascular Surgery 2023; 58: 949–956. 10.1016/j.jvs.2013.04.036

6 Lind B, Morcos O, Ferral H, Chen A, Aquisto T, Lee S, et al. Endovascular Strategies in the Management of Acute Limb Ischemia. Vascular Specialist International 2019; 35(1): 4–9. 10.5758/vsi.2019.35.1.4

7 Enezate T, Omran J, Mahmud E, Patel M, Abu-Fadel M, White C, et al. Endovascular versus surgical treatment for acute limb ischaemia: a systematic review and meta-analysis of clinical trials. Cardiovascular Diagnosis and Therapy 2017; 7(3): 264–271. 10.21037/cdt.2017.03.03

8 Darwood R, Berridge D, Kessel D, Robertson I, Forster R. Surgery versus thrombolysis for initial management of acute limb ischaemia. Cochrane Database of Systematic Reviews 2018; 8: CD002784. 10.1002/14651858.CD002784.pub3

9 Karnabatidis D, Spiliopoulos S, Tsetis D, Siablis D. Quality improvement guidelines for percutaneous catheter-directed intra-arterial thrombolysis and mechanical thrombectomy for acute lower-limb ischemia. Cardiovascular Interventional Radiology 2011; 34(6): 1123–1136. 10.1007/s00270-011-0258-z

10. Jarosinski M, Kennedy J, Khamzina Y, Alie-Cusson F, Tzeng E, Eslami M, et al. Percutaneous thrombectomy for acute limb ischemia is associated with equivalent limb and mortality outcomes compared with open thrombectomy. Journal of Vascular Surgery 2024; 79(5): 1151–1162.e3. 10.1016/j.jvs.2024.01.014

11. Becton, Dickinson and Company (2020). Refining Atherectomy: Rotarex™ Rotational Excisional Atherectomy System. https://ncvh.org/wp-content/uploads/2020/11/BD-Refining-Atherectomy-Aspiration-Together.pdf

12. Muli Jogi R, Damodharan K, Leong H, Tan A, Chandramohan S, Venkatanarasimha N, et al. Catheter-directed thrombolysis versus percutaneous mechanical thrombectomy in the management of acute limb ischemia: a single center review. CVIR Endovascular 2018; 1: 35. 10.1186/s42155-018-0041-1

13. Giusca S, Raupp D, Dreyer D, Eisenbach C, Korosoglou G. Successful endovascular treatment in patients with acute thromboembolic ischemia of the lower limb including the crural arteries. World Journal of Cardiology 2018; 10(10): 145 – 152. 10.4330/wjc.v10.i10.145

14. Ontario. Mechanical Thrombectomy for Acute and Subacute Blocked Arteries and Veins in the Lower Limbs: A Health Technology Assessment. Ontario Technology Assessment Series 2023; 23(1): 1–244. https://www.publications.gov.on.ca/CL32555

15. Barker T, Migliavaca C, Stein C, Colpani V, Falavigna M, Aromataris E, et al. Conducting proportional meta-analysis in different types of systematic reviews: a guide for synthesisers of evidence. BMC Medical Research Methodology 2021; 21: 189. 10.1186/s12874-021-01381-z

16. Page, M, McKenzie J, Bossuyt P, et al. The PRISMA 2020 statement: an updated guideline for reporting systematic reviews. Systematic Reviews 2021; 10: 89. 10.1186/s13643-021-01626-4

17. Stanberry B, Monsalve Roquero B. A systematic review and proportional meta-analysis of percutaneous mechanical thrombectomy for acute and subacute limb ischaemia. PROSPERO 2025 CRD420251015846. https://www.crd.york.ac.uk/PROSPERO/view/CRD420251015846

18. Seo H, Kim S, Lee Y, Park J. RoBANS 2: A Revised Risk of Bias Assessment Tool for Nonrandomized Studies of Interventions. Korean Journal of Family Medicine 2023; 44(5): 249 – 260. 10.4082/kjfm.23.0034

19. Sterne J, Hernán M, Reeves B, Savović J, Berkman N, Viswanathan M, et al. ROBINS-I: a tool for assessing risk of bias in non-randomised studies of interventions. BMJ 2016; 355: i4919. 10.1136/bmj.i4919

20. Zhang Y, Huang L, Wang D, Ren P, Hong Q, Kang D. The ROBINS-I and the NOS had similar reliability but differed in applicability: a random sampling observational studies of systematic reviews/meta-analysis. Journal of Evidence-Based Medicine 2021; 14(2): 112 – 122. 10.1111/jebm.12427

21. Thode R, Solanki G, Aggarwal A, Belekar V, Lakkakula U, Goyal R. A comparison of NOS and ROBINS-I tools for quality assessment of observational studies. Value in Health 2021; 24(S1): S169. 10.1016/j.jval.2021.04.838

22. Brennan S, Johnston R. Research Note: Interpreting findings of a systematic review using GRADE methods. Journal of Physiotherapy 2023; 69(3): 198 – 202. 10.1016/j.jphys.2023.05.012

23. Barendregt J, Doi S, Lee Y, Norman R, Vos T. Meta-analysis of prevalence. Journal of Epidemiology and Community Health 2013; 67(11): 974–978. 10.1136/jech-2013-203104

24. Li W, Xing Y, Feng H, Chen X, Zhang Z. Percutaneous mechanical thrombectomy using the Rotarex^®^S device for the treatment of acute lower limb artery embolism: A retrospective single-center, single-arm study. Frontiers in Surgery 2023; 9: 1017045. 10.3389/fsurg.2022.1017045

25. Fan W, Lu S, Tan J, Cui X, Liang K, Zhu L, et al. Midterm Results of Drug-Coated Balloon Alone or Combined with Rotarex Thrombectomy Device for Treatment of Subacute Femoropopliteal Artery Thrombotic Occlusion. Annals of Vascular Surgery 2023; 92: 240 – 248. 10.1016/j.avsg.2022.11.019

26. Artzner C, Martin I, Hefferman G, Artzner K, Lescan M, de Graaf R, et al. Safety and Efficacy of Rotational Thrombectomy for Treatment of Arterial Occlusions of the Lower Extremities: A Large Single-Center Retrospective Study. Röfo 2023; 195(5): 406–415. 10.1055/a-1952-0092

27. Zhao L, Cai H, Song Q. Clinical Study on Treatment of Acute Lower Extremity Arterial Embolism With Straub Thrombus Removal System. Frontiers in Surgery 2022; 9: 891649. 10.3389/fsurg.2022.891649

28. Yang X, Li X, Yin M, Wang R, Ye K, Lu X, et al. Percutaneous Mechanical Thrombectomy for Acute Limb Ischemia With Aorto-iliac Occlusion. Frontiers in Surgery 2022; 9: 831922. 10.3389/fsurg.2022.831922

29. Wang C, Lu C, Hsieh L, Kuo C, Huang P, Chang K, et al. Comparison of pharmaco-mechanical thrombolysis and catheter-directed thrombolysis for treating thrombotic or embolic arterial occlusion of the lower limb. International Angiology 2022; 41(4): 292 – 302. 10.23736/S0392-9590.22.04809-X

30. Liu L, Zhao J, Bi J, Dai X, Zhang X, He J, et al. Percutaneous Mechanical Atherothrombectomy Using the Rotarex Device in Acute Ischemic Disease of Lower Limbs: A China Retrospective Multicenter Study on 186 Patients. Annals of Vascular Surgery 2022; 85: 146 – 155. 10.1016/j.avsg.2022.02.026

31. Stahlberg E, Anton S, Sieren M, Wegner F, Barkhausen J, Goltz J. Mechanical rotational thrombectomy in long femoropopliteal artery and bypass occlusions: risk factors for periprocedural peripheral embolization. Diagnostic and Interventional Radiology 2021; 27(2): 249 – 256. 10.5152/dir.2021.20100

32. Gong M, He X, Zhao B, Kong J, Gu J, Chen G. Endovascular revascularization strategies using catheter-based thrombectomy versus conventional catheter-directed thrombolysis for acute limb ischemia. Thrombus Journal 2021; 19(1): 96. 10.1186/s12959-021-00349-9

33. Fluck F, Stephan M, Augustin A, Rickert N, Bley TA, Kickuth R. Percutaneous mechanical thrombectomy in acute and subacute lower-extremity ischemia: impact of adjunctive, solely nonthrombolytic endovascular procedures. Diagnostic Interventional Radiology 2021; 27(2): 206 – 213. 10.5152/dir.2021.19403

34. Wang Q, Zhu R, Ren H, Leng R, Zhang W, Li C. Combination of Percutaneous Rotational Thrombectomy and Drug-Coated Balloon for Treatment of Femoropopliteal Artery Nonembolic Occlusion: 12-Month Follow-up. Journal of Vascular and Interventional Radiology 2020; 31(10): 1661 – 1667. 10.1016/j.jvir.2020.03.014

35. Migliara B, Cappellari T, Mirandola M, Griso A, Kolasa K, Zah V, et al. Treatment of bypass failure in patients with chronic limb threatening ischemia - open surgery vs. percutaneous mechanical thrombectomy. Vasa 2020; 49(5): 395 – 402. 10.1024/0301-1526/a000883

36. Liu J, Li T, Huang W, Zhao N, Liu H, Zhao H, et al. Percutaneous mechanical thrombectomy using Rotarex catheter in peripheral artery occlusion diseases - Experience from a single center. Vascular 2019; 27(2): 199 – 203. 10.1177/1708538118813239

37. Liang S, Zhou L, Ye K, Lu X. Limb Salvage After Percutaneous Mechanical Thrombectomy in Patients with Acute Lower Limb Ischemia: A Retrospective Analysis from Two Institutions. Annals of Vascular Surgery 2019; 58: 151 – 159. 10.1016/j.avsg.2018.11.025

38. Latacz P, Simka M, Brzegowy P, Piwowarczyk M, Popiela T. Mechanical rotational thrombectomy with Rotarex system augmented with drug-eluting balloon angioplasty versus stenting for the treatment of acute thrombotic and critical limb ischaemia in the femoropopliteal segment. Wideochir Inne Tech Maloinwazyjne. 2019; 14(2): 311 – 319. 10.5114/wiitm.2018.80006

39. Bulvas M, Sommerová Z, Vaněk I, Weiss J. Prospective Single-Arm Trial of Endovascular Mechanical Debulking as Initial Therapy in Patients With Acute and Subacute Lower Limb Ischemia: One-Year Outcomes. Journal of Endovascular Therapy 2019; 26(3): 291 – 301. 10.1177/1526602819840697

40. Kronlage M, Printz I, Vogel B, Blessing E, Müller OJ, Katus HA, et al. A comparative study on endovascular treatment of (sub)acute critical limb ischemia: mechanical thrombectomy vs thrombolysis. Drug Design Development and Therapy 2017; 11: 1233 – 1241. 10.2147/DDDT.S131503

41. Heller S, Lubanda J, Varejka P, Chochola M, Prochazka P, Rucka D, et al. Percutaneous Mechanical Thrombectomy Using Rotarex® S Device in Acute Limb Ischemia in Infrainguinal Occlusions. Biomed Research International 2017; 2362769. 10.1155/2017/2362769

42. Freitas B, Steiner S, Bausback Y, Branzan D, Ülrich M, Bräunlich S, et al. Rotarex Mechanical Debulking in Acute and Subacute Arterial Lesions. Angiology 2017; 68(3): 233 – 241. 10.1177/0003319716646682

43. Stanek F, Ouhrabkova R, Prochazka D. Percutaneous mechanical thrombectomy in the treatment of acute and subacute occlusions of the peripheral arteries and bypasses. Vasa 2016; 45(1): 49 – 56. 10.1024/0301-1526/a000495

44. Scheer F, Lüdtke C, Kamusella P, Wiggermann P, Vieweg H, Schlöricke E, et al. Combination of rotational atherothrombectomy and Paclitaxel-coated angioplasty for femoropopliteal occlusion. Clinical Medicine Insights: Cardiology 2015; 8(Suppl 2): 43 – 48. 10.4137/CMC.S15231

45. Wissgott C, Kamusella P, Andresen R. Recanalization of acute and subacute venous and synthetic bypass-graft occlusions with a mechanical rotational catheter. CardioVascular and Interventional Radiology 2013; 36(4): 936 – 942. 10.1007/s00270-012-0507-9

46. Lichtenberg M, Stahlhoff W, Boese D, Hailer B. Twelve months outcome after percutaneous mechanical thrombectomy for treatment of acute femoropopliteal bypass occlusion. Cardiovascular Intervention and Therapeutics 2013; 28(2): 178 – 183. 10.1007/s12928-012-0152-x

47. Hundt W, Kalinowski M, Stamm A, Portig I, Swaid Z, Dietz C, et al. Combined treatment of subacute and acute synthetic and venous bypass-graft occlusions with percutaneous mechanical thrombectomy and thrombolysis. European Journal of Radiology 2013; 82(12): e807 – 815. 10.1016/j.ejrad.2013.08.016

48. Sacks D, McClenny T, Cardella J, Lewis C. Society of Interventional Radiology clinical practice guidelines. Journal of Vascular and Interventional Radiology 2003; 14 (9 Pt 2): S199 – 202. 10.1097/01.rvi.0000094584.83406.3e

49. Conte M, Geraghty P, Bradbury A, Hevelone N, Lipsitz S, Moneta G, et al. Suggested objective performance goals and clinical trial design for evaluating catheter-based treatment of critical limb ischemia. Journal of Vascular Surgery 2009; 50(6): 1462 – 1473. e1-3. 10.1016/j.jvs.2009.09.044

50. Loffroy R, Falvo N, Galland C, Fréchier L, Ledan F, Midulla M, et al. Percutaneous Rotational Mechanical Atherectomy Plus Thrombectomy Using Rotarex S Device in Patients With Acute and Subacute Lower Limb Ischemia: A Review of Safety, Efficacy, and Outcomes. Frontiers in Cardiovascular Medicine 2020; 7: 557420. 10.3389/fcvm.2020.557420

51. Ebben H, Jongkind V, Wisselink W, Hoksbergen A, Yeung K. Catheter Directed Thrombolysis Protocols for Peripheral Arterial Occlusions: a Systematic Review. European Journal of Vascular and Endovascular Surgery 2019; 57(5): 667 – 675. 10.1016/j.ejvs.2018.11.018

52. Results of a prospective randomized trial evaluating surgery versus thrombolysis for ischemia of the lower extremity. The STILE trial. Annals of Surgery 1994; 220(3): 251 – 66. 10.1097/00000658-199409000-00003

53. Ouriel K, Veith F, Sasahara A. Thrombolysis or peripheral arterial surgery: phase I results. TOPAS Investigators. Journal of Vascular Surgery 1996; 23(1): 64 – 73. 10.1016/s0741-5214(05)80036-9

54. Ouriel K, Shortell C, DeWeese J, Green R, Francis C, Azodo M. A comparison of thrombolytic therapy with operative revascularization in the initial treatment of acute peripheral arterial ischemia. Journal of Vascular Surgery 1994; 19(6): 1021 – 1030. 10.1016/s0741-5214(94)70214-4

55. de Donato G, Setacci F, Sirignano P, Galzerano G, Massaroni R, Setacci C. The combination of surgical embolectomy and endovascular techniques may improve outcomes of patients with acute lower limb ischemia. Journal of Vascular Surgery 2014; 59(3): 729 – 736. 10.1016/j.jvs.2013.09.016

56. Doelare S, Koedam T, Ebben H, Tournoij E, Hoksbergen A, Yeung K, et al. Catheter Directed Thrombolysis for Not Immediately Threatening Acute Limb Ischaemia: Systematic Review and Meta-Analysis. European Journal of Vascular and Endovascular Surgery 2023; 65(4): 537 – 545. 10.1016/j.ejvs.2022.12.030

57. Wang J, Kim A, Kashyap V. Open surgical or endovascular revascularization for acute limb ischemia. Journal of Vascular Surgery 2016; 63(1): 270 – 278. 10.1016/j.jvs.2015.09.055

58. Stanberry B, Maclean D, Elbasty A. Percutaneous mechanical atherothrombectomy versus arterial bypass surgery for femoropopliteal in-stent restenosis: a budget impact analysis. Journal of the Society for Cardiovascular Angiography and Interventions 2025; 4(6): 103616. 10.1016/j.jscai.2025.103616

59. Higgins J, Thomas J, Chandler J, Cumpston M, Li T, Page M, et al, editor(s). Cochrane Handbook for Systematic Reviews of Interventions. 2nd Edition. Chichester (UK): John Wiley & Sons, 2019. https://www.cochrane.org/authors/handbooks-and-manuals/handbook/current

