## Supplemental Tables for "Rotational Percutaneous Mechanical Thrombectomy for Acute and Subacute Limb Ischaemia: a Systematic Review and Proportional Meta-analysis"

| Supplementary Table S1. PRISMA 2020 Checklist | | | |
|---|---|---|---|
| **Topic** | **No.** | **Item** | **Location where item is reported** |
| **TITLE** | | | |
| **Title** | 1 | Identify the report as a systematic review. | Page 1, lines 1 - 4. |
| **ABSTRACT** | | | |
| **Abstract** | 2 | See the PRISMA 2020 for Abstracts checklist below | Pages 1 – 2, lines 22 - 43. |
| **TITLE** | | | |
| **Title** | A1 | Identify the report as a systematic review. | Yes |
| **BACKGROUND** | | | |
| **Objectives** | A2 | Provide an explicit statement of the main objective(s) or question(s) the review addresses. | Yes |
| **METHODS** | | | |
| **Eligibility criteria** | A3 | Specify the inclusion and exclusion criteria for the review. | Yes |
| **Information sources** | A4 | Specify the information sources (e.g. databases, registers) used to identify studies and the date when each was last searched. | Yes |
| **Risk of bias** | A5 | Specify the methods used to assess risk of bias in the included studies. | Yes |
| **Synthesis of results** | A6 | Specify the methods used to present and synthesize results. | Yes |
| **RESULTS** | | | |
| **Included studies** | A7 | Give the total number of included studies and participants and summarise relevant characteristics of studies. | Yes |
| **Synthesis of results** | A8 | Present results for main outcomes, preferably indicating the number of included studies and participants for each. If meta-analysis was done, report the summary estimate and confidence/credible interval. If comparing groups, indicate the direction of the effect (i.e. which group is favoured). | Yes |
| **DISCUSSION** | | | |
| **Limitations of evidence** | A9 | Provide a brief summary of the limitations of the evidence included in the review (e.g. study risk of bias, inconsistency and imprecision). | Yes |
| **Interpretation** | A10 | Provide a general interpretation of the results and important implications. | Yes |
| **OTHER** | | | |
| **Funding** | A11 | Specify the primary source of funding for the review. | Yes |
| **Registration** | A12 | Provide the register name and registration number. | Yes |
| **INTRODUCTION** | | | |

| Supplementary Table S1. PRISMA 2020 Checklist | | | |
|---|---|---|---|
| **Topic** | **No.** | **Item** | **Location where item is reported** |
| **Rationale** | 3 | Describe the rationale for the review in the context of existing knowledge. | Page 4, lines 115 - 117. |
| **Objectives** | 4 | Provide an explicit statement of the objective(s) or question(s) the review addresses. | Page 4, lines 117 - 121. |
| **METHODS** | | | |
| **Eligibility criteria** | 5 | Specify the inclusion and exclusion criteria for the review and how studies were grouped for the syntheses. | Page 5, lines 128 - 138. |
| **Information sources** | 6 | Specify all databases, registers, websites, organisations, reference lists and other sources searched or consulted to identify studies. Specify the date when each source was last searched or consulted. | Page 5, lines 140 - 144. |
| **Search strategy** | 7 | Present the full search strategies for all databases, registers and websites, including any filters and limits used. | Page 5, lines 145 - 153. |
| **Selection process** | 8 | Specify the methods used to decide whether a study met the inclusion criteria of the review, including how many reviewers screened each record and each report retrieved, whether they worked independently, and if applicable, details of automation tools used in the process. | Pages 5 - 6, lines 153 - 156. |
| **Data collection process** | 9 | Specify the methods used to collect data from reports, including how many reviewers collected data from each report, whether they worked independently, any processes for obtaining or confirming data from study investigators, and if applicable, details of automation tools used in the process. | Page 7, lines 158 - 163. |
| **Data items** | 10a | List and define all outcomes for which data were sought. Specify whether all results that were compatible with each outcome domain in each study were sought (e.g. for all measures, time points, analyses), and if not, the methods used to decide which results to collect. | Page 7, lines 163 - 167. |
| | 10b | List and define all other variables for which data were sought (e.g. participant and intervention characteristics, funding sources). Describe any assumptions made about any missing or unclear information. | Page 7, lines 158 - 162. |
| **Study risk of bias assessment** | 11 | Specify the methods used to assess risk of bias in the included studies, including details of the tool(s) used, how many reviewers assessed each study and whether they worked independently, and if applicable, details of automation tools used in the process. | Page 7, lines 169 - 174. |
| **Effect measures** | 12 | Specify for each outcome the effect measure(s) (e.g. risk ratio, mean difference) used in the synthesis or presentation of results. | Page 7, lines 163 - 167. |
| **Synthesis methods** | 13a | Describe the processes used to decide which studies were eligible for each synthesis (e.g. tabulating the study intervention characteristics and comparing against the planned groups for each synthesis (item 5)). | Page 7, lines 178 - 183. |
| | 13b | Describe any methods required to prepare the data for presentation or synthesis, such as handling of missing summary statistics, or data conversions. | Page 7, lines 178 - 189. |

| Supplementary Table S1. PRISMA 2020 Checklist | | | |
|---|---|---|---|
| **Topic** | **No.** | **Item** | **Location where item is reported** |
| | 13c | Describe any methods used to tabulate or visually display results of individual studies and syntheses. | Page 7, lines 186 - 187. |
| | 13d | Describe any methods used to synthesize results and provide a rationale for the choice(s). If meta-analysis was performed, describe the model(s), method(s) to identify the presence and extent of statistical heterogeneity, and software package(s) used. | Page 7, lines 183 - 187. |
| | 13e | Describe any methods used to explore possible causes of heterogeneity among study results (e.g. subgroup analysis, meta-regression). | Page 7, line 185. |
| | 13f | Describe any sensitivity analyses conducted to assess robustness of the synthesized results. | Page 7, line 189. |
| **Reporting bias assessment** | 14 | Describe any methods used to assess risk of bias due to missing results in a synthesis (arising from reporting biases). | Page 7, line 169 - 173. |
| **Certainty assessment** | 15 | Describe any methods used to assess certainty (or confidence) in the body of evidence for an outcome. | Page 7, lines 174 - 176 |
| **RESULTS** | | | |
| **Study selection** | 16a | Describe the results of the search and selection process, from the number of records identified in the search to the number of studies included in the review, ideally using a flow diagram. | Page 8, lines 192 - 197. |
| | 16b | Cite studies that might appear to meet the inclusion criteria, but which were excluded, and explain why they were excluded. | Page 8, lines 192 - 197. |
| **Study characteristics** | 17 | Cite each included study and present its characteristics. | Page 9, lines 199 - 218. |
| **Risk of bias in studies** | 18 | Present assessments of risk of bias for each included study. | Page 10, lines 234 - 236. Supplementary Table S3. |
| **Results of individual studies** | 19 | For all outcomes, present, for each study: (a) summary statistics for each group (where appropriate) and (b) an effect estimate and its precision (e.g. confidence/credible interval), ideally using structured tables or plots. | Tables 2 & 3, pages 14 - 18. |
| **Results of syntheses** | 20a | For each synthesis, briefly summarise the characteristics and risk of bias among contributing studies. | Table 4, page 19. |
| | 20b | Present results of all statistical syntheses conducted. If meta-analysis was done, present for each the summary estimate and its precision (e.g. confidence/credible interval) and measures of statistical heterogeneity. If comparing groups, describe the direction of the effect. | Pages 10 - 13, lines 237 - 357. |
| | 20c | Present results of all investigations of possible causes of heterogeneity among study results. | Table 4, page 19. |
| | 20d | Present results of all sensitivity analyses conducted to assess the robustness of the synthesized results. | Page 13, lines 345 - 346. |
| **Reporting biases** | 21 | Present assessments of risk of bias due to missing results (arising from reporting biases) for each synthesis assessed. | Page 10, lines 234 - 246. Supplementary Table S3. |
| **Certainty of evidence** | 22 | Present assessments of certainty (or confidence) in the body of evidence for each outcome assessed. | Table 4, page 19. |

| Supplementary Table S1. PRISMA 2020 Checklist | | | |
|---|---|---|---|
| **Topic** | **No.** | **Item** | **Location where item is reported** |
| **DISCUSSION** | | | |
| **Discussion** | 23a | Provide a general interpretation of the results in the context of other evidence. | Pages 27 - 28, lines 367 - 401. |
| | 23b | Discuss any limitations of the evidence included in the review. | Page 28, lines 412 - 429. |
| | 23c | Discuss any limitations of the review processes used. | Page 28, lines 412 - 429. |
| | 23d | Discuss implications of the results for practice, policy, and future research. | Page 28, lines 403 - 412. |
| **OTHER INFORMATION** | | | |
| **Registration and protocol** | 24a | Provide registration information for the review, including register name and registration number, or state that the review was not registered. | Page 5, lines 123 - 126. |
| | 24b | Indicate where the review protocol can be accessed, or state that a protocol was not prepared. | Page 5, lines 123 - 126. Reference 17. |
| | 24c | Describe and explain any amendments to information provided at registration or in the protocol. | Page 5, lines 133 - 134. |
| **Support** | 25 | Describe sources of financial or non-financial support for the review, and the role of the funders or sponsors in the review. | Page 2, lines 45 - 47. |
| **Competing interests** | 26 | Declare any competing interests of review authors. | Pages 2 - 3, lines 48 - 70 |
| **Availability of data, code and other materials** | 27 | Report which of the following are publicly available and where they can be found: template data collection forms; data extracted from included studies; data used for all analyses; analytic code; any other materials used in the review. | Tables 2 - 3, pages 14 - 18. Supplementary Tables. |
| **ABSTRACT** | | | |

| Supplementary Table S2. **Search strategies** | | |
|---|---|---|
| **Databases (Dialog):** MEDLINE® (search strategy run and results downloaded on 7 November 2024) | | |
| **Set#** | **Searched for** | **Results** |
| S1 | Rotarex | 93 |
| S2 | MESH.EXACT("Mechanical Thrombolysis") | 1058 |
| S3 | (MESH.EXACT("Atherectomy") OR MESH.EXACT("Thrombectomy")) and ti,ab,if(mechanical or rotational or rotation) | 4206 |
| S4 | ti,ab,if((mechanical or rotational or rotation) near/5 (atherectom* or atheroablat* or thrombectom*)) or ti,ab,if(rotablat* or "mechanical thrombolysis") | 9101 |
| S5 | MESH.EXACT("Lower Extremity") OR MESH.EXACT("Leg") OR MESH.EXACT("Chronic Limb-Threatening Ischemia") OR MESH.EXACT("Limb Salvage") OR (MESH.EXACT("Femoral Artery") and MESH.EXACT("Popliteal Artery")) OR MESH.EXACT("Peripheral Arterial Disease") | 111423 |
| S6 | ti,ab,if(limb or limbs or leg or legs or "lower extremity" or "lower extremities" or (femoral and popliteal) or femoropopliteal or "femoro-popliteal" or crural or "acute extremit*" or "subacute extremit*" or "critical extremit*") | 432090 |
| S7 | ti,ab,if(peripheral near/3 (arterial or artery or arteries or ischaemi* or ischemi*)) | 39455 |
| **S8** | **s1 and pd(1999-2024)** | **93** |
| **Databases (Dialog):** Embase®, Embase preprints (search strategy run and results downloaded on 7 November 2024) | | |
| **Set#** | **Searched for** | **Results** |
| S1 | rotarex | 251 |
| S2 | EMB.EXACT("rotational atherectomy") OR EMB.EXACT.EXPLODE("rotational atherectomy device") OR EMB.EXACT("mechanical thrombectomy") OR EMB.EXACT("rotational thrombectomy catheter") | 18041 |
| S3 | (EMB.EXACT("atherectomy") OR EMB.EXACT("atherectomy catheter") OR EMB.EXACT("atherectomy device") OR EMB.EXACT("percutaneous thrombectomy") OR EMB.EXACT("thrombectomy catheter") OR EMB.EXACT("thrombectomy device") OR EMB.EXACT("surgical thrombectomy") OR EMB.EXACT("thrombectomy")) and ti,ab,if(mechanical or rotational or rotation) | 6253 |
| S4 | ti,ab,if((mechanical or rotational or rotation) near/5 (atherectom* or atheroablat* or thrombectom*)) or ti,ab,if(rotablat* or "mechanical thrombolysis") | 16237 |
| S5 | EMB.EXACT("limb ischemia") OR EMB.EXACT("acute limb ischemia") OR EMB.EXACT("critical limb ischemia") OR EMB.EXACT("lower limb") OR EMB.EXACT("limb blood vessel") OR EMB.EXACT("leg") OR EMB.EXACT("leg blood vessel") OR EMB.EXACT("limb") OR EMB.EXACT("limb blood flow") OR EMB.EXACT("leg disease") OR EMB.EXACT("limb disease") OR EMB.EXACT("limb salvage") OR EMB.EXACT("leg artery") OR EMB.EXACT("leg ischemia") OR EMB.EXACT("peripheral arterial disease") OR EMB.EXACT("peripheral arterial occlusion") OR EMB.EXACT("peripheral occlusive artery disease") OR EMB.EXACT("peripheral ischemia") | 362209 |
| S6 | (EMB.EXACT.EXPLODE("femoral artery") OR EMB.EXACT("femoral artery flow") OR EMB.EXACT.EXPLODE("femoral artery stent")) and (EMB.EXACT("popliteal artery") OR EMB.EXACT("popliteal artery stent")) | 4542 |

| | | |
|---|---|---|
| S7 | ti,ab,if(limb or limbs or leg or legs or "lower extremity" or "lower extremities" or (femoral and popliteal) or femoropopliteal or "femoro-popliteal" or crural or "acute extremit*" or "subacute extremit*" or "critical extremit*") | 636300 |
| S8 | ti,ab,if(peripheral near/3 (arterial or artery or arteries or ischaemi* or ischemi*)) | 63022 |
| **S9** | **s1 and pd(1999-2024)** | **251** |

| **Databases (Dialog):** Cochrane Library® (search strategy run and results downloaded on 7 November 2024) | | |
|---|---|---|
| **ID#** | **Searched for** | **Results** |
| #1 | (rotarex):ti,ab,kw | 5 |
| #2 | MeSH descriptor: [Mechanical Thrombolysis] this term only | 75 |
| #3 | MeSH descriptor: [Atherectomy] this term only | 59 |
| #4 | MeSH descriptor: [Thrombectomy] this term only | 747 |
| #5 | (mechanical or rotational or rotation):ti,ab,kw | 42,152 |
| #6 | (#3 or #4) and #5 | 241 |
| #7 | ((mechanical or rotational or rotation) near/5 (atherectom* or atheroablat* or thrombectom*)):ti,ab,kw | 1053 |
| #8 | (rotablat* or "mechanical thrombolysis"):ti,ab,kw | 146 |
| #9 | #1 or #2 or #6 or #7 or #8 | 1154 |
| #10 | MeSH descriptor: [Lower Extremity] this term only | 2248 |
| #11 | MeSH descriptor: [Leg] this term only | 3695 |
| #12 | MeSH descriptor: [Chronic Limb-Threatening Ischemia] this term only | 60 |
| #13 | MeSH descriptor: [Limb Salvage] this term only | 182 |
| #14 | MeSH descriptor: [Femoral Artery] this term only | 1417 |
| #15 | MeSH descriptor: [Popliteal Artery] this term only | 571 |
| #16 | #14 and #15 | 444 |
| #17 | MeSH descriptor: [Peripheral Arterial Disease] this term only | 1789 |
| #18 | (limb or limbs or leg or legs or "lower extremity" or "lower extremities" or (femoral and popliteal) or femoropopliteal or "femoro-popliteal" or crural or (acute next extremit*) or (subacute next extremit*) or (critical next extremit*)):ti,ab,kw | 70565 |
| #19 | (peripheral near/3 (arterial or artery or arteries or ischaemi* or ischemi*)):ti,ab,kw | 6967 |
| #20 | #10 or #11 or #12 or #13 or #16 or #17 or #18 or #19 | 74951 |
| #21 | #9 and #20 | 74 |
| #22 | **#1 and publication date range: 1999-2024** | **5** |

**Supplementary Table S3.** **RoBANS2 risk of bias assessment**

| Study | Comparability of Target Group | Target Group Selection | Confounders | Measurement of Intervention | Blinding of Assessors | Outcome Assessment | Incomplete Outcome Data | Selective Outcome Reporting | Overall Risk |
|---|---|---|---|---|---|---|---|---|---|
| Li, 2023 [24] | Low | High | High | Low | Low | Low | Low | High | **High** |
| Fan, 2023 [25] | Low | High | High | Low | Low | Low | Low | High | **High** |
| Artzner, 2022 [26] | Low | High | High | Low | Low | Low | Low | High | **High** |
| Zhao, 2022 [27] | Low | High | High | Low | Low | Low | Low | High | **High** |
| Yang, 2022 [28] | Low | High | High | Low | Low | Low | Unclear | High | **High** |
| Wang, 2022 [29] | Low | Low | High | Low | Low | Low | Low | High | **Moderate** |
| Liu et al, 2022 [30] | Unclear | High | High | Low | Low | Low | Low | High | **High** |
| Stahlberg, 2021 [31] | Low | High | High | Low | Low | Low | Low | High | **High** |
| Gong, 2021 [33] | Low | High | High | Low | Low | Low | Low | High | **High** |
| Fluck, 2021 [33] | Low | High | High | Low | Unclear | Low | Low | High | **High** |
| Wang, 2020 [34] | Low | High | High | Low | Low | Low | Low | High | **High** |
| Migliara, 2020 [35] | Low | High | High | Low | Low | Low | Low | High | **High** |
| Liu, 2019 [36] | Low | High | High | Low | Low | Low | Low | High | **High** |
| Liang, 2019 [37] | Low | High | High | Low | Low | Low | Low | High | **High** |
| Latacz, 2019 [38] | Low | High | High | Low | Low | Low | Low | High | **High** |
| Bulvas, 2019 [39] | Low | Low | High | Low | Low | Low | Low | High | **Moderate** |
| Kronlage, 2017 [40] | Low | High | High | Low | Low | Low | Unclear | High | **High** |

| Supplementary Table S3. RoBANS2 risk of bias assessment | | | | | | | | | |
|---|---|---|---|---|---|---|---|---|---|
| **Study** | **Comparability of Target Group** | **Target Group Selection** | **Confounders** | **Measurement of Intervention** | **Blinding of Assessors** | **Outcome Assessment** | **Incomplete Outcome Data** | **Selective Outcome Reporting** | **Overall Risk** |
| Heller, 2017 [41] | Low | High | High | Low | Low | Low | Low | High | **High** |
| Freitas, 2017 [42] | Low | High | High | Low | Low | Unclear | Low | High | **High** |
| Stanek, 2016 [43] | Low | Low | High | Low | Low | Low | Low | High | **Moderate** |
| Scheer, 2015 [44] | Low | High | High | Low | Low | Low | Low | High | **High** |
| Wissgott, 2013 [45] | Low | High | High | Low | Low | Low | Low | High | **High** |
| Lichtenberg, 2013 [46] | Low | High | High | Low | Unclear | Low | Low | High | **High** |
| Hundt et al, 2013 [47] | Low | High | High | Low | Low | Low | Low | High | **High** |
